# Longitudinal entomological study using molecular xenomonitoring of *Wuchereria bancrofti* in *Culex quinquefasciatus* in three Nepalese districts: Implications for lymphatic filariasis transmission assessment and elimination

**DOI:** 10.64898/2026.10.23.26366133

**Authors:** Deena Shrestha, Bijay Bajracharya, Pooja Shah, Padma Shrestha, Sushmita Ghimire, Satya Raj Paudel, Swornima Dangol, Pramod Shrestha, Shiva Rajbanshi, Parakrami Ranjitkar, Sangeeta Chand, Meena Sharma, Chandra Kishore Sardar, Rojita Tuladhar, Sushila Bajracharya, Ajit Kumar Karna, Krishna Das Manadhar, Ram Kumar Mahato, Gokarna Dahal, Tijana Williams, Louise A. Kelly Hope

**Affiliations:** Center for Health and Disease Studies, Kathmandu, Nepal; Department of Biotechnology, Tribhuvan University, Kirtipur, Kathmandu, Nepal; Government of Nepal Ministry of Health and Population, Department of Health Services, Kathmandu, Epidemiology and Disease Control Division, Nepal; GSK, London, UK; Institute of Infection, Veterinary and Ecological Sciences, University of Liverpool, UK

## Abstract

**Background:** In Nepal, lymphatic filariasis (LF), caused by *Wuchereria bancrofti*, remains a priority neglected tropical disease targeted for elimination by 2030. Despite large-scale mass drug administration (MDA) and post-MDA surveillance, focal transmission may persist. Molecular xenomonitoring (MX) of mosquito vectors is a complementary method for detecting ongoing transmission. This study assessed mosquito abundance and species composition and compared gravid traps and CDC light traps for capturing *Culex quinquefasciatus*, the primary LF vector. It examined spatial and seasonal vector patterns, *W. bancrofti* infection rates, and climatic associations across three LF endemic districts.

**Methodology:** A longitudinal entomological survey was conducted from September 2022 to September 2023 in selected sites of Dhading, Jhapa, and Kapilvastu districts using gravid traps and CDC light traps. Mosquitoes were morphologically identified to species level, and female *Cx. quinquefasciatus* were pooled (up to 25 mosquitoes) for detection of *W. bancrofti* DNA using real-time PCR. Mosquito abundance, species composition, trap efficiency, spatial variation (within and between locations), temporal trends (daily, weekly, monthly, seasonal), infection rate (pool positivity, and maximum likelihood estimate (MLE) of infection using PoolTestR package in R software), and climatic associations (precipitation, temperature, humidity) were analysed.

**Results:** A total of 60,922 mosquitoes belonging to eight genera (*Culex, Anopheles, Aedes, Armigeres, Malaya, Mansonia, Tripteroides,* and *Toxorhynchites*) were identified. Of these, 16,044 were female *Cx. quinquefasciatus*. Gravid traps showed higher relative trapping efficiency of female *Cx. quinquefasciatus* than light traps, with the highest efficiency observed in Jhapa (3.60:1). Temporal trends varied across locations; however, overall *Cx. quinquefasciatus* abundance peaked during the spring and summer months (March–May), coinciding with the monsoon period, and was positively correlated with climatic factors. *W. bancrofti* infection rates varied by location and trap type, with gravid traps yielding higher infection rates, particularly in Jhapa. A total of 785 pools of *Cx. quinquefasciatus* were screened. Pool positivity and infection prevalence (MLE) ranged from 10.3% to 17.6% and 0.5% to 0.9% in Dhading, 12.5% to 36.8% and 0.9% to 2.0% in Jhapa, and 5.3% to 20.0% and 0.3% to 1.5% in Kapilvastu, respectively.

**Conclusions:** The study highlights ongoing transmission in three LF-endemic areas of Nepal, emphasising the need for continued surveillance in high-risk areas and periods. Gravid trap–based xenomonitoring could complement community-based *W. bancrofti* surveillance by identifying hotspots and optimising the timing and targeting of LF elimination interventions.

## Introduction

Lymphatic filariasis (LF) is one of the World Health Organization (WHO)’s priority neglected tropical diseases (NTDs), targeted for elimination by 2030 as set out in the NTD road map 2021−2030 [1]. Globally, there are currently 856x million people at risk of LF across more than xxx endemic countries. LF is a parasitic disease caused predominately by microscopic, thread-like worms called *Wuchereria bancrofti*. The disease spreads from person-to-person through bites on an infected mosquito belonging to the genera *Culex*. Aedes and *Anopheles* [2].

To eliminate filarial worms from the human body and prevent subsequent transmission, the main strategy is preventive chemotherapy using a two-or three-drug regimen delivered through three to five annual rounds of mass drug administration (MDA) [3,4]. To assess interruption of transmission and whether microfilariae prevalence has fallen below the critical threshold, the WHO recommends Transmission Assessment Surveys (TAS), which are conducted three times. If TAS results fail to fall below the critical threshold, endemic districts or implementation units are required to repeat MDA. These surveys can be a setback for programmes and are often costly. Therefore, additional surveillance approaches and adaptive targeted strategies focusing on hotspot or high-risk areas are needed.

Monitoring vector abundance and parasite infection rates in vectors is one such option and can be a useful approach to evaluating transmission [5,6], and assessing the potential for integrating surveillance into vector control programmes [7]. Molecular xenomonitoring (MX), which refers to the detection of parasite DNA in mosquitoes [2] has been used to track infection pre- and post-MDA [8], and in post-MDA and validation phases [8–12]. Preliminary threshold levels of infection prevalence—less than 0.25% for *Culex* spp., less than 1% for *Anopheles* spp., and less than 0.1% for *Aedes* spp.—are recommended to determine whether MDA can be stopped [2].

LF is widespread throughout the South-East Asian region [13,14]. In Nepal, LF is a considerable public health problem across 61 of the 77 districts [15,16]. The Nepal National LF Programme has conducted prevalence mapping using the immunochromatographic test (ICT) to detect circulating *W. bancrofti* antigens in 2001 and 2005, with remapping in 2012. Prevalence rates were found to range between 0 to 39 with infection detected across different topographical areas ranging in altitude from 100 m above sea level in the lowland Terai ecological zone to 1700 m above sea level in Hill areas [16]. Comparatively, more LF cases are reported in the Terai than in the Hills; however, valleys and river basin areas of hilly districts also have evidence of infection and morbidity cases. LF is more prevalent in rural areas, predominantly affecting poorer communities. *W. bancrofti* is the only recorded parasite in Nepal, and the mosquito *Culex quinquefasciatus* is considered the main vector responsible for transmission, with gravid traps identified as useful surveillance tools [17–21].

In 2021, the Nepal LF programme had made substantial progress in scaling up MDA, and several districts have passed TAS and stopped MDA. Dhading District was one of the first districts to stop MDA, and after completion of the three TAS rounds, no further routine surveillance was conducted, although there was evidence of residual positivity [21,22] and many morbidity cases found in assessment studies [23]. In addition, some districts in the highly endemic Terai region had failed TAS and required further rounds of MDA and additional approached to assess the transmission risk areas.

To strengthen surveillance and assess the potential for MX, this study aimed to conduct entomological assessments and evaluate two trapping types (gravid trap, CDC light trap), across three distinct locations in Nepal: one site in a previously endemic district that had stopped MDA (Dhading District), and two sites in districts with ongoing MDA and failed TAS (Jhapa and Kapilvastu Districts).

The specific objectives were to:

1. Characterize mosquito abundance and species composition.
2. Compare *Cx.* abundance and gravidity by trap type (gravid trap, CDC light trap), specifically focus on *Cx. quinquefasciatus* distributions.
3. Examine fine-scale spatial variation in *Cx. quinquefasciatus* abundance within each location, exploring differences between trap type and local environmental characteristics.
4. Examine temporal and seasonal patterns of *Cx. quinquefasciatus* abundance and associated climatic factors.
5. Determine the spatial and temporal distribution of *W. bancrofti* infection in *Cx. quinquefasciatus* mosquitoes and associated climatic factors.

## Methods

### Study design and site characteristics

Longitudinal entomological collections were conducted over a 6 to 13-month period in three locations of the country with different ecological characteristics and stages of LF programme elimination. The study was reported in accordance with the Strengthening the reporting of observational studies in epidemiology (STROBE) guidelines for cohort studies (see S1 checklist).

Vector collections were obtained using two trapping methods. The study was designed to assess the spatial – temporal patterns of mosquito species distribution and abundance, with a specific focus on *Cx. quinquefasciatus* and their *W. bancrofti* infection rates.

Study locations included one village in each of Dhading, Jhapa, and Kapilvastu districts (Fig 1), representing Hill and Terai ecological zones of Nepal, which differ in altitude, climate, land use and can affect vector distributions [24]. Fig 1 map was created using ArcGIS 10.7 (ESRI, Redland, CA, USA), with administrative boundaries sourced from Hermes download center [25], and elevation data based on CGIAR SRTM 90m Digital Elevation Databases [26]. A summary of the study location is presented in Table 1.

**Figure 1.**
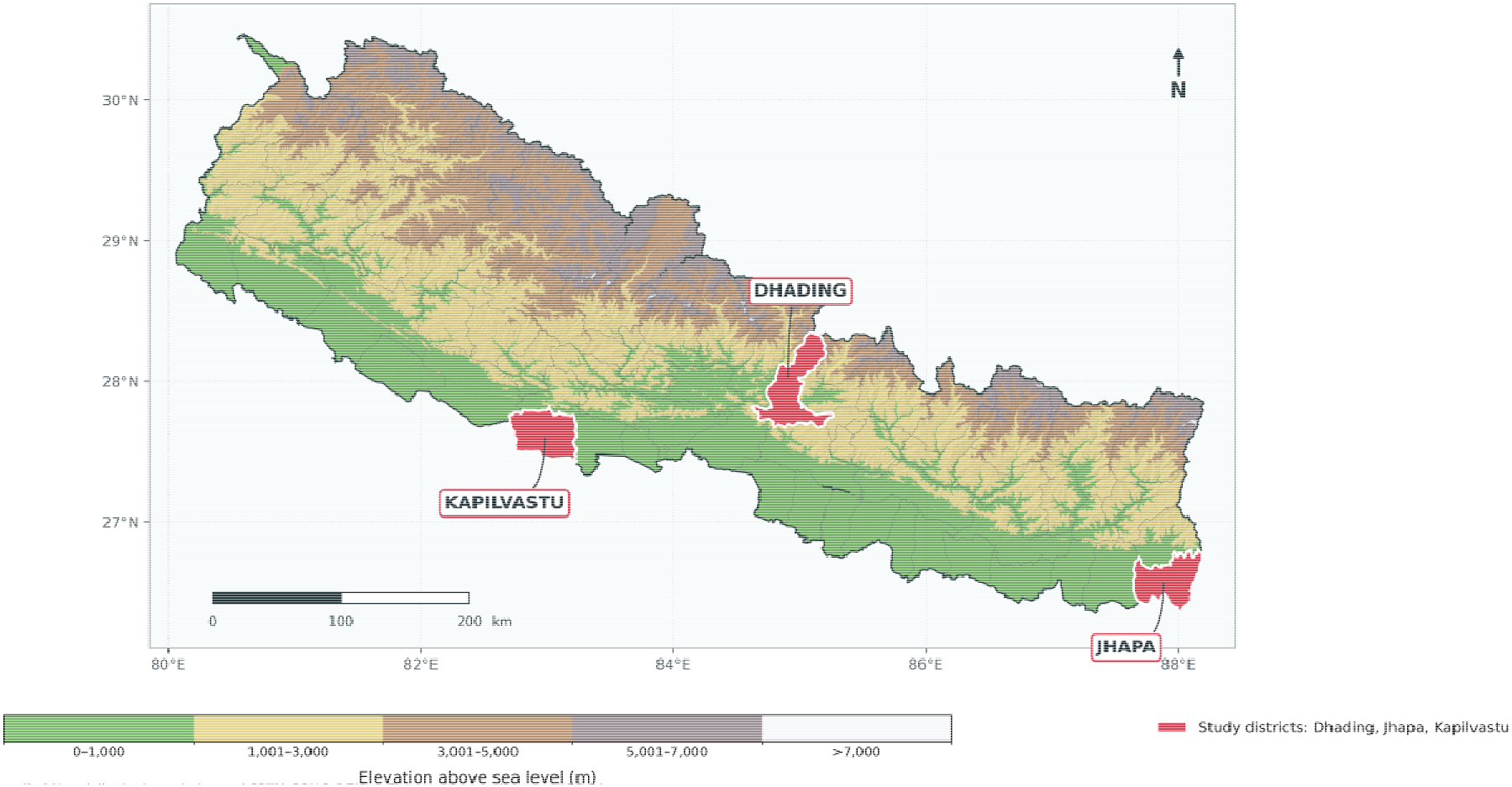
Map showing study districts and elevation.

**Table 1.**
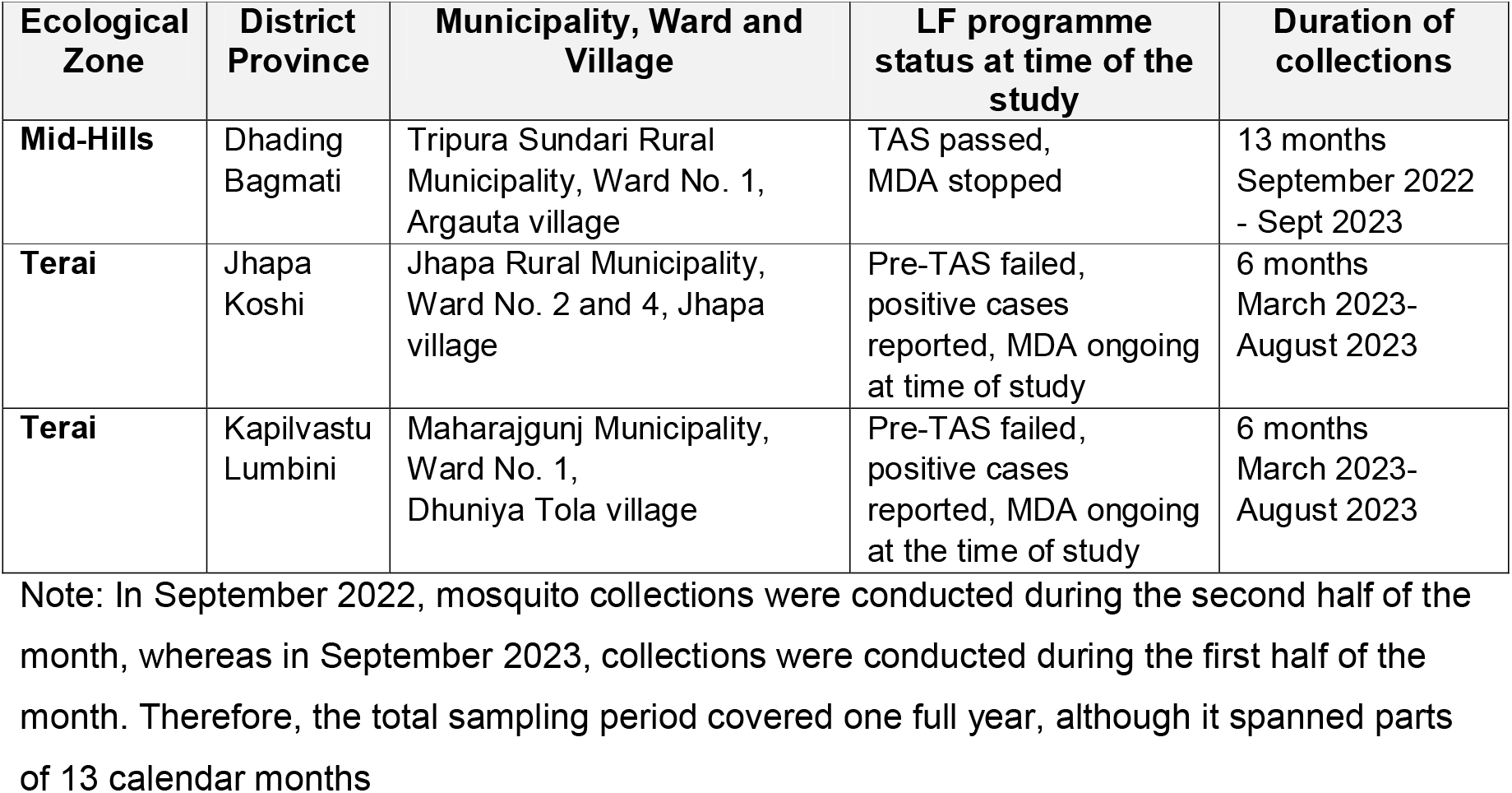
Summary of study locations characteristics.

In Dhading District, entomological collections were conducted for 13 months in Argauta village, Tripura Sundari Rural Municipality (Table 1). This location was purposively selected as MDA was stopped in 2013 and post-MDA surveillance (TAS 1, TAS 2 and TAS 3) has been completed by 2018. Recent human positivity has been detected since stopping MDA [21,22], along with a high number of LF morbidity cases identified through house-to-house patient mapping [23]. The village was selected for the study as a hotspot area within the district.

In Jhapa District (Eastern Nepal) and Kapilvastu District (Western Nepal) entomological collections were conducted in six months of the year in Jhapa village and Dhuniya Tola village, respectively (Table 1). These districts were selected due to repeated failed Pre-TAS, and the specific villages were selected based on reported positive cases during Pre-TAS [27–29]. The study period focused on the expected peak transmission period and began in the pre-monsoon period in last week of March 2023. The collections focused on the potential peak monsoon period due to funding limitations.

### Placement of gravid traps and light traps

Gravid traps and CDC light traps [30] were used for collecting mosquitoes primarily in two different sites within each village. Traps were placed approximately 500 meters apart. The sites were selected based on differing housing types, presence of LF morbidity cases, potential mosquito breeding habitats, and the feasibility of trap installation, to capture a range of local environmental conditions and potential risk factors for transmission [14]. Based on these criteria Site 1 and Site 2 were allocated within the study site. In Jhapa and Kapilvastu locations an additional trap was set up in a third site (Site 3) for a short time to maximise collections while the opportunity was available. A summary of the site characteristics and trap types in the three study locations is provided in S1 file.

### Mosquito collection, identification and recording

Mosquitoes were primarily collected and identified daily (5-7 days per week) by two experienced entomologists at each study location. Mosquito genera identification was conducted using the morphological keys [17]. The sex (male, female) and the physiological status (gravid) of each female mosquito were recorded. Only intact mosquitoes were used. Once identified, mosquitoes were placed in 2 ml/1.5 ml Eppendorf tubes with 70% ethanol, and data on species were recorded on paper sheets and then entered into Microsoft Excel (Microsoft Corporation, Redmond, WA, USA) before being sent to the central laboratory in Kathmandu.

### Molecular screening of mosquitoes for *W. bancrofti* infection

In the central laboratory, mosquito samples were divided into pools of maximum 25 mosquitoes by species, location, site and time of collection. After removing the wings and legs of mosquitoes, DNA was extracted from each pool of mosquitoes using the DNeasy Tissue Kit (Qiagen, Valencia, California, USA) following the manufacturer’s instructions. DNA quality was measured prior to PCR using a NanoDrop (Thermo Fisher Scientific, USA) [31].

Pooled mosquitoes were tested for the presence of *W. bancrofti* DNA based on Rao et al., 2006 [32]. PCR amplification was performed using *W. bancrofti* specific primers LDR1 and LDR2 for real-time detection. Pools of mosquitoes were tested for *W. bancrofti* DNA by TaqMan real-time PCR. The sequence of primers and probes used, and specific procedure, is provided in S1 file.

### Data analyses

All collected data were entered into Microsoft Excel (Microsoft Corporation, Redmond, WA, USA) spreadsheets, then cleaned and manually edited as required. Data analysis was conducted using SPSS version 31.0 (IBM Corp., Armonk, NY, USA). Descriptive analyses were performed primarily based on monthly data collated from the daily and epidemiological week data. Epidemiological weeks are widely used in disease surveillance systems to standardise the reporting and help comparisons across space and time [33,34]. Specific statistical tests were conducted in relation to each study objective and are described here. The level of statistical significance was set at *p* < 0.05.

Objective 1.

- To characterize mosquito abundance and species composition from the three study locations mosquitoes were identified as described above and data summarised by main genera including *Culex* spp. *Armigeres* spp. *Anopheles* spp. *Aedes* spp. *Mansonia* spp. and Malaya spp.

Objective 2.

- To compare mosquito abundance between traps at each location, data were summarised and statistical difference examined using a Mann–Whitney U test. The relative trapping efficiency of gravid traps to light traps was determined by calculating the ratios.

Objective 3.

- To explore the fine-scale distributions of *Cx. quinquefasciatus* that may be associated with trap type and local environmental characteristics, only data from each gravid trap and light trap at the two main sites (Site 1 and Site 2) were summarised. The overall differences between each site in the number of *Cx. quinquefasciatus was* determined using a Mann–Whitney U test.

Objective 4.

- To determine temporal (monthly, weekly, daily) and seasonal patterns of *Cx. quinquefasciatus* abundance and associated climatic factors were analysed. Data from each location over the 6 to13-month collection period were summarised. Time-specific comparisons of the daily mean numbers of female *Cx. quinquefasciatus* by trap type and month were conducted using data collected from Site 1 and Site 2 over the 6-month (March to August 2023) for each location.
**-** Monthly climatic factors including precipitation, temperature and humidity, were examined in relation to monthly daily mean *Cx. quinquefasciatus* abundance using Pearson’s correlation analysis. Precipitation (total millimetre (mm)) data were obtained from CHIRPS high resolution rainfall estimates from rain gauge and satellite observations [35,36]. Temperature (average °C) and humidity (average %) data were collected from/obtained from digital thermo-hygrometer).

Objective 5. Infection poolscreen

- To determine spatial and temporal distribution of *W. bancrofti* infection in *Cx. quinquefasciatus* mosquitoes and associated climatic factors each mosquito pool was tested by PCR for filarial parasite DNA as described above. Pools grouped by epidemiological weeks and classified as positive if at least one mosquito was infected and the pool positivity for each epidemiological week calculated. *W. bancrofti* DNA Infection prevalence in *Cx. quinquefasciatus* was estimated using the Maximum Likelihood Estimates (MLE) with 95% Confidence Intervals, calculated with the PoolTestR package in R software [37,38].
- Analyses focused on (i) onsite × trap combinations and (ii) monthly trends based on combined weekly infection data within each study location to assess spatial - temporal variation. Due to limited sample sizes, some finer-scale analyses were not conducted such as monthly comparison by trap type.

### Ethical consideration

The project was approved by the Social Welfare Council prior to implementation and once the protocols were finalized in accordance with government rules. Ethical approval for the study was obtained from the Ethical Review Board of the Nepal Veterinary Research Council (Ref: 209/2078/2079).

## Results

### Objective 1. Characterize mosquito abundance and species composition

A total of 60,922 mosquitoes were collected from the three district locations: Dhading (n = 26,996), Jhapa (n = 17,060), and Kapilvastu (n = 16,866). Altogether eight genera of mosquitoes (*Culex, Anopheles, Aedes, Armigeres, Malaya, Mansonia, Tripteroides and Toxorhynchites*) were collected by the different traps across the study period. A summary of the data is presented in Table 2 and Fig 2. Specific species information at each location and trap type is provided in S1 file.

**Figure 2.**
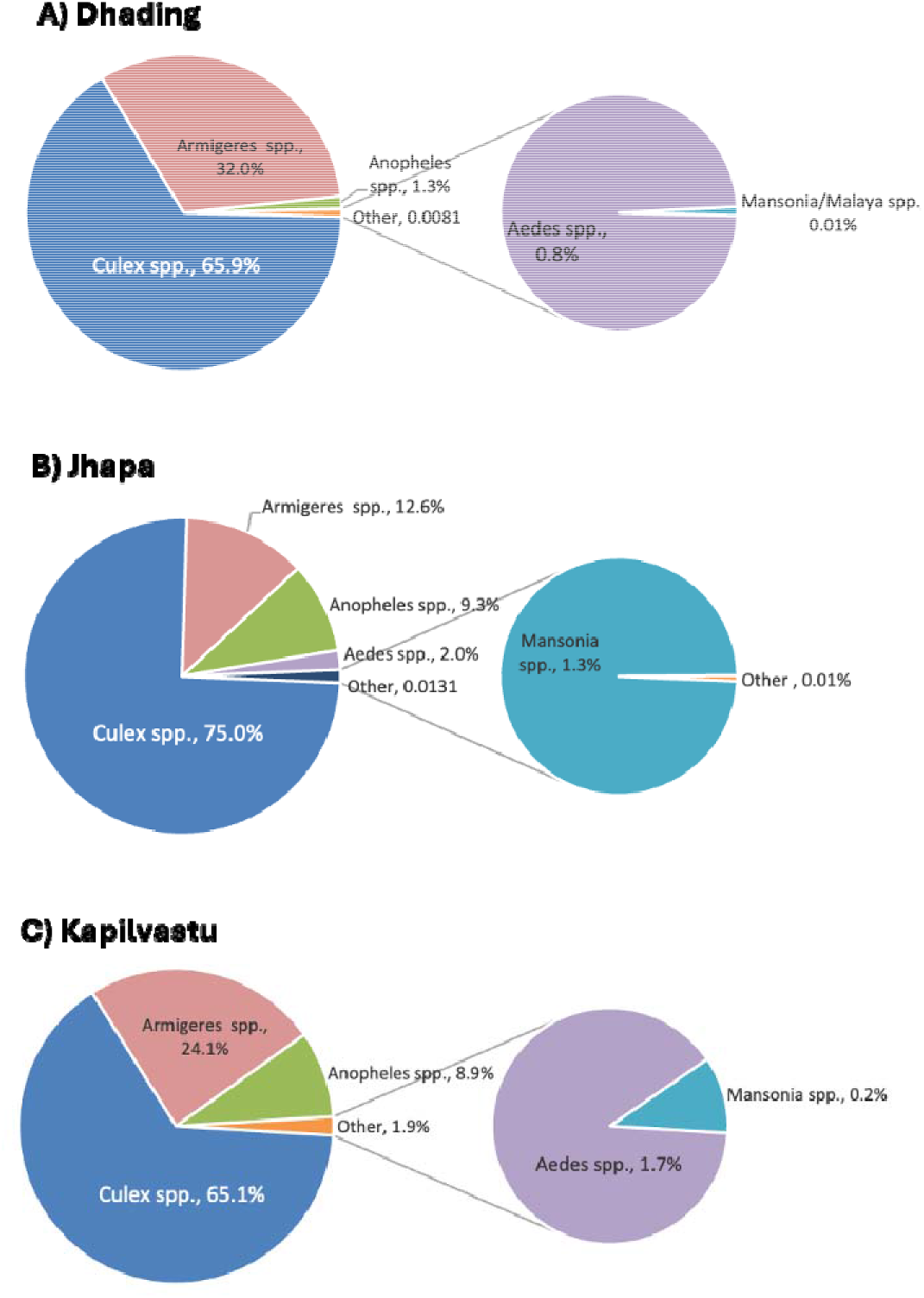
Graphical presentation of main mosquito genera in three district locations.

**Table 2.**
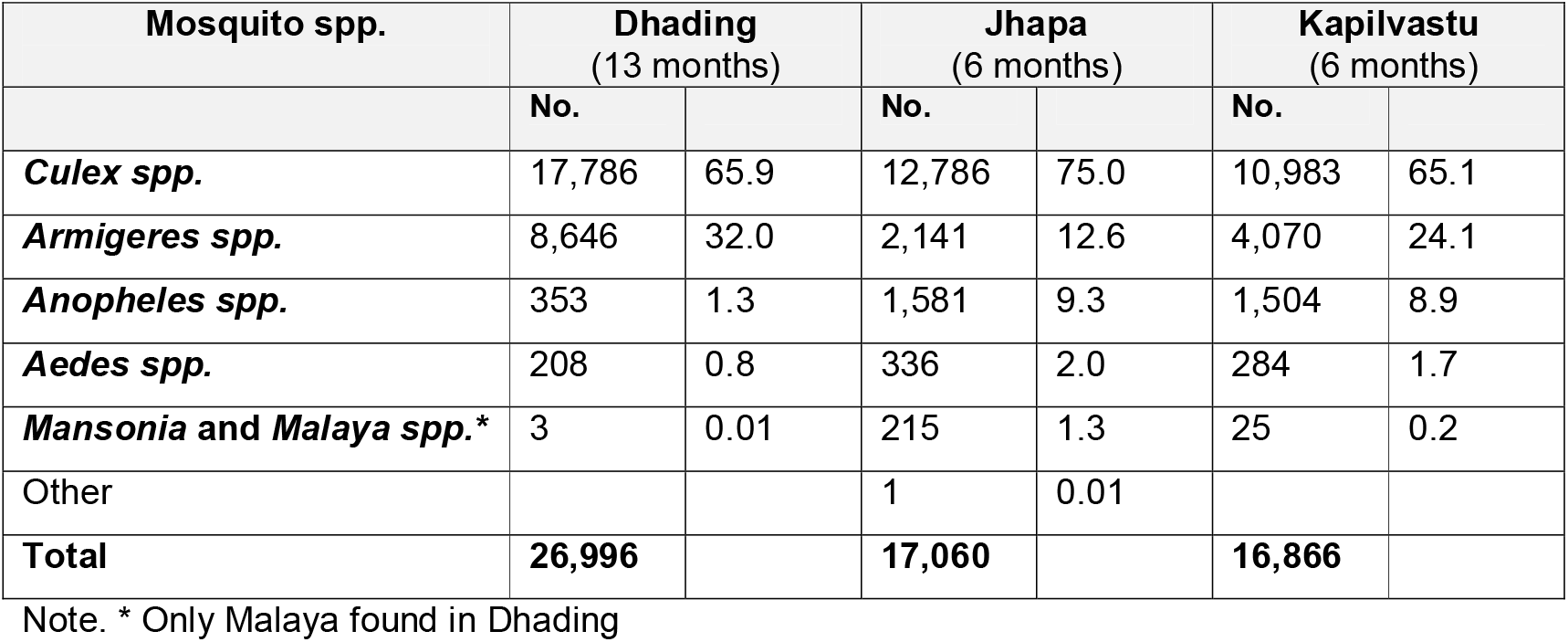
Summary of mosquito genera and abundance in three district locations.

In Dhading, collections were conducted from two trap sites within the village from September 2022 to September 2023. *Culex spp*. (n = 17,786, 65.9%) were most abundant, followed by *Armigeres spp*. (n = 8,646, 32.0%). *Anopheles spp*. (n = 353, 1.3%) and *Aedes spp*. (n = 208, 0.8%) were comparatively rare, while *Mansonia spp.* and *Malaya spp*. were negligible (n = 3; 0.01%).

In Jhapa, collections were conducted from two trap sites within the village from March to August 2023, and one additional site from July to August 2023 to boost the collection during the monsoon period. *Culex spp*. was most abundant (n = 12,786, 75.0%), followed by *Armigeres spp*. (n = 2,141, 12.6%) and *Anopheles spp*. (n = 1,581. 9.3%). *Aedes spp*. (n = 336, 2.0%) and *Mansonia spp*. (n = 215, 1.3%) were also recorded, with a very small “other” category ( n = 1. 0.01%).

In Kapilvastu, collections were conducted from two sites within the village from March to August 2023, and one additional site in August 2023 to boost the collection during monsoon period. *Culex spp*. (n = 10,983, 65.1%) were most abundant, followed by *Armigeres spp*. which represented around one quarter of collections (n = 4,070, 24.1%,). *Anopheles spp*. (n = 1,504, 8.9%), *Mansonia spp*. (n = 284, 1.7%), and *Aedes spp*. (n = 25, 0.15%) comprised smaller proportions.

### Objective 2. Compare *Cx. quinquefasciatus* abundance and gravidity by trap type

#### Abundance

The number of mosquitoes, and specifically, the number of female *Cx. quinquefasciatus* varied markedly by trap type and district locations. A summary of the data is presented in Table 3 and detailed information is provided in S1 file.

**Table 3.** Comparison of trap type numbers and efficiency for female *Culex quinquefasciatus* for Site 1 and Site 2 in each location.

| District<br>(collection<br>period) | Trap<br>Type | Total No.<br>Mosquitoes<br>Collected | Female *Cx. quinquefasciatus* |  |  | Mann-Whitney U<br>p-value |
| --- | --- | --- | --- | --- | --- | --- |
|  |  |  | No. | Percentage<br>of total | Relative<br>Trapping<br>Efficiency<br>(Gravid to Light) |  |
| **Dhading**<br>(13<br>months) | Light | 17,410 | 3,271 | 18.8 | 1.19: 1 | p<0.001** |
|  | Gravid | 9,586 | 3,894 | 40.6 |  |  |
| **Jhapa**<br>(6 months) | Light | 9,083 | 1,132 | 12.5 | 3.60: 1 | p<0.001** |
|  | Gravid | 7,977 | 4,076 | 51.1 |  |  |
| **Kapilvastu**<br>(6 months) | Light | 9,463 | 1,342 | 14.2 | 1.74: 1 | p<0.001** |
|  | Gravid | 7,403 | 2,329 | 31.5 |  |  |

**Note.** Gravid traps placed outdoors and light traps placed indoors. Note that only Site 1 and 2 from each location were included in the analysis. In Dhading, Site 1 light trap was placed in a cow shed, whereas Site 2 was placed indoor. Statistical significance set at \* $p<0.05$ and \*\* $p<0.001$ .

Overall, light traps (n=35,956; 59.0%) collected a greater number of mosquitoes than gravid traps (n=24,966; 40.1%). Conversely, gravid traps (n=10,299; 64.2%) collected nearly double the number of female *Cx. quinquefasciatus* than light traps (n=5,745; 35.8%). In Dhading, Jhapa and Kapilvastu gravid traps represented 40.6%, 51.1% and 31.5% of all female *Cx. quinquefasciatus* mosquitoes collected respectively, which were significantly higher than light traps (p<0.001). The relative trapping efficiency of gravid traps to light traps for female *Cx. quinquefasciatus* was highest in Jhapa (3.60 :1), followed by Kapilvastu (1.74: 1) and Dhading (1.19: 1) (Table 3).

#### Gravidity

A higher proportion of gravid *Cx. quinquefasciatus* were found in gravid traps than light traps across all districts (p<0.001). In Dhading, gravid traps (80.1%) collected approximately 3.8 times more gravid *Cx. quinquefasciatus* than light traps (20.9%); in Jhapa 3.5 times more (gravid trap=50.8%; light trap=14.4%), and in Kapilvastu, 1.4 times more (gravid trap=47.9%; light trap=34.2%). See S1 file for details.

### Objective 3. Site and trap specific *Cx. quinquefasciatus* distributions within each location

For each locations, data from the gravid trap and light trap at the two main sites (Site 1 and Site 2) over the 6-month period (March to August 2023) included the numbers of *Cx. quinquefasciatus* mosquitoes (females and males). These were compared with the combined abundance of all other mosquito species.

Overall, the abundance patterns of *Cx. quinquefasciatus* differed significantly between Site 1 and Site 2 in each study location. Gravid traps consistently captured higher numbers than light traps, despite variation in site characteristics. Fig 3 provides a graphical summary of the results, and corresponding data are tabulated in S1 file.

**Figure 3.**
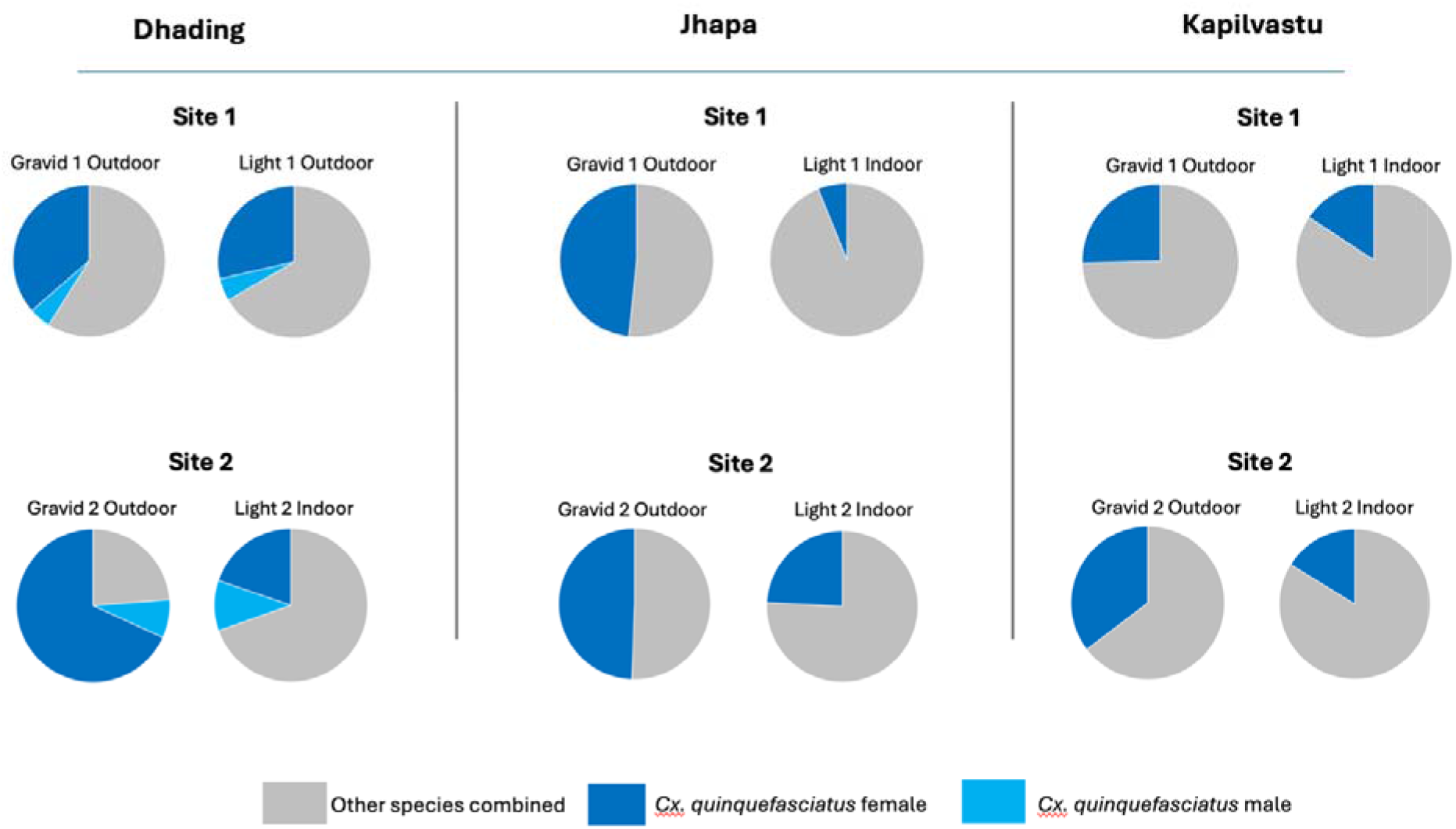
Graphical presentation of *Cx. quinquefasciatus* and other mosquito species abundance by location, site and trap type.

In Dhading, overall, there were significant differences in the number of *Cx. quinquefasciatus* between sites, with Site 2 yielding higher numbers than Site 1 (Mann-Whitney U = 666780; p<0.001). Site 1 consisted predominantly of concrete houses, cattle were present within household compounds, and no LF morbidity cases reported. Female *Cx. quinquefasciatus* comprised 36.4% (n=1,240/3,406) of collections by gravid traps and 28.6% (n=576/2,012) of collections by light traps. In contrast, Site 2 comprised houses constructed mainly of mud and wood, cattle were present within household compounds, and LF morbidity cases were reported from this site. Female *Cx. quinquefasciatus* comprised 68.2% (n=1,828/2,681) of collections by gravid traps and 19.8% (n=2,320/11,739) of collections by light traps. Overall, there was a small number and proportion of male *Cx. quinquefasciatus* found in all sites.

In Jhapa, overall, there were significant differences in the number of *Cx. quinquefasciatus* between sites, with Site 2 yielding higher numbers than Site 1 (Mann-Whitney U = 16776; p<0.031). Site 1 consisted of a concrete house where field officers stay with no LF morbidity case in the house. Female *Cx. quinquefasciatus* comprised 48.3% (n=1,603/3,316) of collections by gravid traps and 6.1% (n=222/3,613) of collections by light traps. In contrast, Site 2 consisted of a dense houses which were made up of wood and mud roofed with galvanized sheets and thatched straw and LF morbidity cases were reported in the site. Female *Cx. quinquefasciatus* comprised 49.4% (n=1,583/3,202) of collections by gravid traps and 24.2% (n=781/3,229) of collections by light traps.

In Kapilvastu, overall, there were significant differences in the number of *Cx. quinquefasciatus* between sites, with Site 2 yielding higher numbers than Site 1 (Mann-Whitney U = 8743; p<0.001). Site 1 consisted of a concrete health centre with no livestock or LF morbidity cases reported. Female *Cx. quinquefasciatus* comprised 25.4% (n=470/1,854) of collections by gravid traps and 15.7% (n=323/2,060) of collections by light traps. Site 2 consisted of predominantly single-story houses constructed of mud, wood, or cement-bonded bricks, with galvanized sheet roofs. The houses were surrounded by gardens containing diverse vegetation, and LF morbidity case had been reported in the area. Female *Cx. quinquefasciatus* comprised 35.4% (n=1,686/4,768) of collections by gravid traps and 16.2% (n=939/5,808) of collections by light traps. No male *Cx. quinquefasciatus* were found.

### Objective 4. Temporal and seasonal patterns of *Cx. quinquefasciatus* abundance and associated climatic factors

#### Monthly and weekly abundance

The weekly total and daily average (mean) number of female *Cx. quinquefasciatus* collected across three study locations varied by season and trap type. Fig 4A graphically presents the weekly data collected over the 6-13-month collection period, and Figs 4B-D present daily mean number of female *Cx. quinquefasciatus* for the 6-month period (March to August 2023) to allow comparisons between study locations. Related data are tabulated in S1 file.

**Figure 4.**
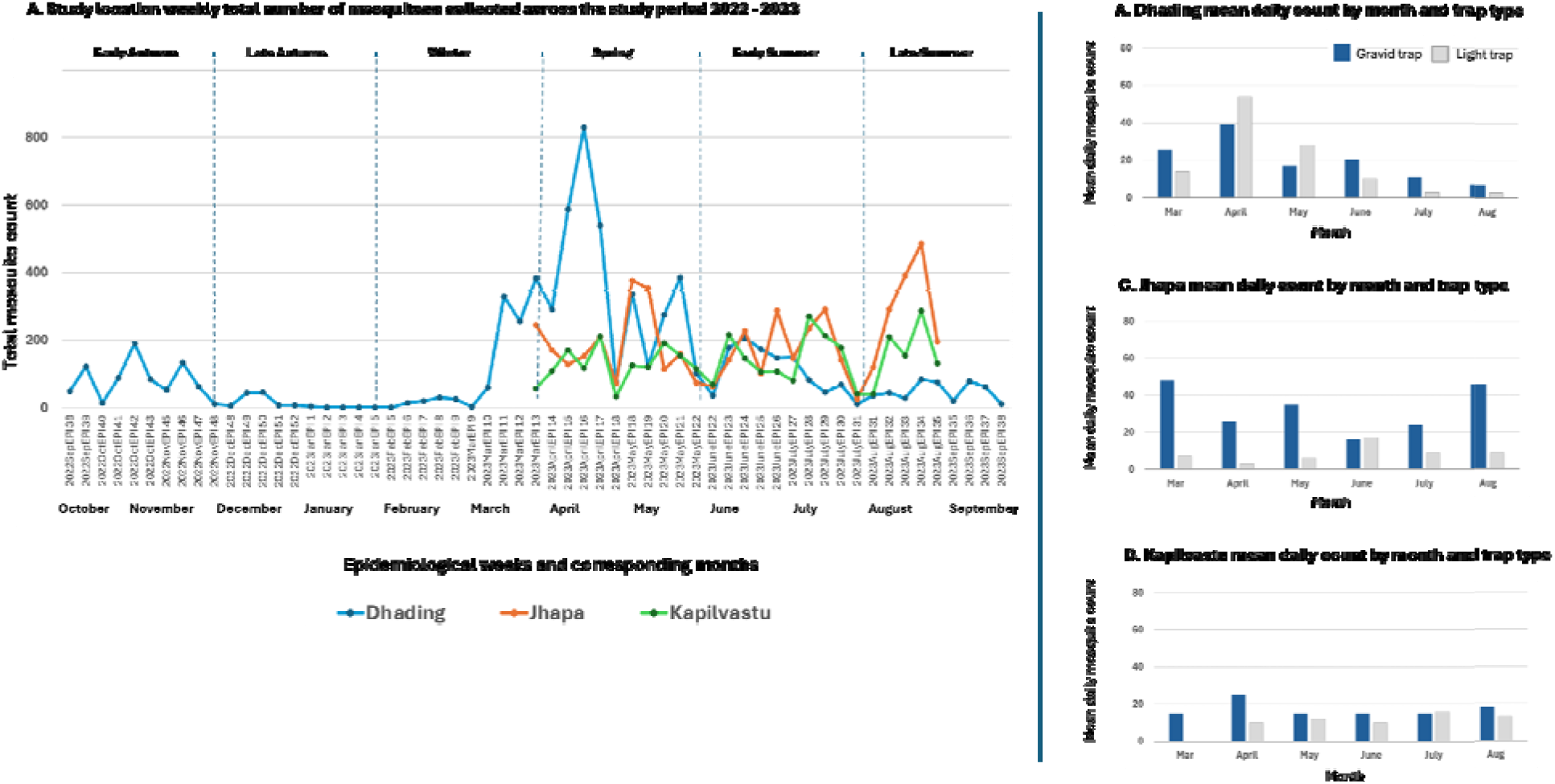
Graphical presentation of weekly total and daily average (mean) number of female *Cx. quinquefasciatus* collected across three study locations.

In Dhading, the temporal analysis showed clear seasonal variation in female *Cx. quinquefasciatus* abundance over the 13-month collection period (Fig 4A). The lowest weekly counts occurred in late autumn and winter, with five or fewer mosquitoes recorded in January 2023 (epidemiological weeks 1–5). In contrast, the highest abundance was observed in spring, with marked peaks during March to May 2023 (epidemiological weeks 11–21), when weekly counts ranged from 77 to 829 mosquitoes.

In Jhapa and Kapilvastu, over the 6-month collection period, the numbers of female *Cx. quinquefasciatus* were relatively stable, with the highest counts observed during summer. The highest weekly mosquito counts were recorded in Jhapa in May (n = 378; epidemiological week 18) and August (n = 485; epidemiological week 34), and in Kapilvastu in July (n = 270; epidemiological week 28) and August (n = 287; epidemiological week 34) (Fig 4A).

### Note that only data from Site 1 and 2 from each location were included in the analysis

#### Daily mean abundance

The daily mean abundance (and daily range) of female *Cx. quinquefasciatus* by trap type and month are tabulated in Table 4 and graphically presented in Fig 4B-D. Overall, peak daily mosquito abundance was observed during the spring and summer months of 2023 across each study location.

**Table 4.** Summary of daily mean abundance of female *Cx. quinquefasciatus,* infection prevalence and monthly climate measures.

| District | Mosquito measures |  |  | Infection prevalence |  | Climate factors |  |  |
| --- | --- | --- | --- | --- | --- | --- | --- | --- |
|  | Gravid trap Daily mean (range) | Light trap Daily mean (range) | Total mosquitoes Daily mean | Pool positivity (%) | MLE prev. (%) | Precip. total (mm) | Temp. average (°C) | Humidity average (%) |
| **Dhading** |  |  |  |  |  |  |  |  |
| Sept-22 | 3 (1-6) | 13 (3-50) | 16 | 10.0 | 0.5 | 92.7 | 24.6 | 68.8 |
| Oct-22 | 17 (2-24) | 6 (2-15) | 21 | 33.3 | 2.0 | 133.1 | 21.2 | 65.1 |
| Nov-22 | 10 (1-48) | 3 (1-5) | 11 | 33.3 | 2.2 | 0.7 | 19.7 | 63.7 |
| Dec-22 | 4 (0-10) | 2 (0-4) | 5 | 0.0 | 0.0 | 8.0 | 15.0 | 78.5 |
| Jan-23 | 1 (0-2) | 0 (0-0) | 1 | 0.0 | 0.0 | 7.6 | 13.1 | 78.1 |
| Feb-23 | 3 (0-8) | 1 (0-4) | 4 | 0.0 | 0.0 | 8.5 | 15.5 | 72.4 |
| Mar-23 | 26 (1-71) | 14 (0-44) | 38 | 8.5 | 0.4 | 53.2 | 18.0 | 67.9 |
| Apr-23 | 39 (1-75) | 54 (6-161) | 93 | 12.4 | 0.6 | 30.8 | 21.2 | 59.4 |
| May-23 | 17 (2-50) | 28 (2-21) | 45 | 17.7 | 0.9 | 126.7 | 22.2 | 77.3 |
| Jun-23 | 20 (1-53) | 10 (1-65) | 29 | 11.1 | 0.6 | 201.1 | 26.2 | 73.9 |
| Jul-23 | 11(2-36) | 3 (1-9) | 14 | 3.8 | 0.3 | 348.8 | 26.8 | 83.8 |
| Aug-23 | 7 (1-22) | 3 (1-9) | 10 | 20.0 | 1.8 | 408.5 | 25.5 | 86.8 |
| Sep-23 | 9 (3-31) | 4 91-9) | 12 | 30.0 | 2.4 | 166.0 | 26.2 | 83.6 |
| **Jhapa** |  |  |  |  |  |  |  |  |
| Mar-23 | 48 (30-73) | 7 (0.7) | 48 | 9.1 | 0.4 | 13.0 | 28.5 | 31.2 |
| Apr-23 | 26 (3-68) | 3 (1-7) | 29 | 18.2 | 1.0 | 36.6 | 31.5 | 32.9 |
| May-23 | 35 (2-123) | 6 (1-73) | 40 | 26.8 | 1.5 | 111.3 | 32.2 | 31.6 |
| Jun-23 | 16 (1-59) | 17 (1-159) | 32 | 18.9 | 1.0 | 228.1 | 32.6 | 42.7 |
| Jul-23 | 24 (1-75) | 9 (1-79) | 32 | 36.7 | 2.4 | 706.2 | 32.4 | 47.0 |
| Aug-23 | 46 (5-141) | 9 (1-34) | 55 | 26.8 | 1.5 | 576.8 | 32.1 | 47.2 |
| **Kapilvastu** |  |  |  |  |  |  |  |  |
| Mar-23 | 23 (7-20) | 0 (0-0) | 23 | 33.3 | 1.9 | 6.9 | 28.1 | 34.4 |
| Apr-23 | 25 (5-48) | 10 (8-11) | 26 | 18.5 | 0.9 | 26.0 | 30.2 | 33.1 |
| May-23 | 15 (4-39) | 12 (3-22) | 26 | 10.3 | 0.5 | 76.4 | 30.9 | 39.2 |
| Jun-23 | 15 (4-32) | 10 (1-20) | 25 | 7.4 | 0.4 | 202.9 | 34.1 | 40.5 |
| Jul-23 | 15 (1-33) | 16 (2-53) | 30 | 9.5 | 0.5 | 367.0 | 31.3 | 60.2 |
| Aug-23 | 18 (3-46) | 13 (3-34) | 31 | 15.0 | 0.8 | 6.9 | 28.1 | 34.4 |

Note: MLE prev. (%) = maximum likelihood estimate prevalence

In Dhading, the daily mean number of female *Cx. quinquefasciatus* was lowest during January and increased from 4 per night in February to 38 per night in March, reaching a peak of 93 per night in April 2023, followed by a gradual decline to September (Table 4). The daily mean number of mosquitoes varied by trap type as shown in Fig 4B. For the period March-August 2023, the daily mean number of *Cx. quinquefasciatus* collected by gravid traps was lowest in August (n = 7) and highest in April (n = 39). Similarly, comparable trends were recorded for light traps with the lowest recorded in July and August (n = 3) and highest in April (n = 54) (Fig 4B).

In Jhapa and Kapilvastu, the daily mean number of female *Cx. quinquefasciatus* remained relatively steady between March and August 2023. In both locations, gravid traps collected more female *Cx. quinquefasciatus* than light traps (Figs 4C-D), with the highest numbers recorded in Jhapa in March (n = 48 and in Kapilvastu in April (n = 25). In contrast, the daily mean number collected by light traps was highest in Jhapa in June (n = 17) and in Kapilvastu in July (n = 16).

#### Temporal and climate analysis

Monthly climatic variables -precipitation (total mm), temperature (average °C), and humidity (average %) - are summarised in Table 4. Correlations for the 6-month period March to August 2023 between climate variables and the daily mean abundance of female *Cx. quinquefasciatus* by trap type showed variable results across each study location (details in S1 file).

In Dhading, correlations between the overall monthly daily mean abundance of female *Cx. quinquefasciatus* and climate factors showed a statistically significant negative relationships with precipitation (r=-0.823; p=0.044) and humidity (r=-0.90; p<0.015), and a negative relationship with temperature (r=-0.573; p=0.235). Similar significant relationships were observed with gravid traps but not light traps (see S1 file for details).

Different trends were observed in Jhapa and Kapilvastu study locations. In Jhapa, a weak positive relationship was observed between overall monthly daily mean female *Cx. quinquefasciatus* and precipitation (r=0.157; p=0.766) and humidity (r=0.084; p=0.874), while a negative relationship was observed with temperature (r=-0.394; p=0.439). These relationships differed by trap type, and none were statistically significant.

In contrast in Kapilvastu, positive relationships were observed between the overall monthly daily mean abundance of female *Cx. quinquefasciatus* and precipitation (r=0.756; p=0.082), temperature (r=0.460; p=0.358) and humidity (r=0.728; p=0.101). Similar relationships were observed with light traps but not gravid traps and none statistically significant (see S1 file for details).

### Objective 5. Infection

#### Site and trap-specific analysis

Overall, a total of 785 pools of female *Cx. quinquefasciatus* mosquitoes were made from all three districts; 357 pools from Dhading, 250 from Jhapa and 178 from Kapilvastu. The pooled PCR analysis showed pool positivity (proportion of PCR-positive pools) varied between and within study locations; Dhading 14.3%, Jhapa 25.6% and Kapilvastu 12.4%. Overall, the infection prevalence (estimated using MLE) in *Cx. quinquefasciatus* (estimated proportion of mosquitoes infected) were low to moderate (Table 5). Pool positivity and infection prevalence (MLE) ranged from 10.3–17.6% and 0.5–0.9% in Dhading, 12.5 36.8% and 0.9–2.0% in Jhapa, and 5.3–20.0% and 0.3–1.5% in Kapilvastu, respectively. Data are summarised in Table 5 and provided in detail in S1 file.

**Table 5.** Pool-based PCR infection estimates of *Wuchereria bancrofti* by district, site, and trap type.

| District | Site | Trap type | Mosquitoes tested (n) | Pools tested (n) | Positive pools (n) | Pool positivity (%) | MLE prevalence | 95% CI |
| --- | --- | --- | --- | --- | --- | --- | --- | --- |
| Dhading | 1 | Gravid | 1,605 | 78 | 8 | 10.3% | 0.5% | 0.2 - 1.0 |
| Dhading | 1 | Light | 626 | 42 | 3 | 7.1% | 0.5% | 0.1- 1.3 |
| Dhading | 2 | Gravid | 2,281 | 112 | 18 | 16.1% | 0.9% | 0.5 - 1.3 |
| Dhading | 2 | Light | 2,635 | 125 | 22 | 17.6% | 0.9% | 0.6 -1.3 |
| Jhapa | 1 | Gravid | 1,603 | 74 | 17 | 23.0% | 1.2% | 0.7 - 1.9 |
| Jhapa | 1 | Light | 222 | 17 | 3 | 17.7% | 1.5% | 0.4 - 3.8 |
| Jhapa | 2 | Gravid | 1,579 | 71 | 20 | 28.2% | 1.5% | 0.9 - 2.2 |
| Jhapa | 2 | Light | 781 | 42 | 9 | 21.4% | 1.3% | 0.6 - 2.4 |
| Jhapa | 3 | Gravid | 890 | 38 | 14 | 36.8% | 2.0% | 1.1 - 3.2 |
| Jhapa | 3 | Light | 129 | 8 | 1 | 12.5% | 0.9% | 0.0 - 3.8 |
| Kapilvastu | 1 | Gravid | 470 | 25 | 3 | 12.0% | 0.7% | 0.2 - 1.7 |
| Kapilvastu | 1 | Light | 323 | 19 | 1 | 5.3% | 0.3% | 0.0 - 1.4 |
| Kapilvastu | 2 | Gravid | 1,686 | 77 | 12 | 15.6% | 0.8% | 0.4 - 1.3 |
| Kapilvastu | 2 | Light | 939 | 45 | 4 | 8.9% | 0.4% | 0.1 - 1.0 |
| Kapilvastu | 3 | Gravid | 173 | 7 | 1 | 14.3% | 0.6% | 0.0 - 2.7 |
| Kapilvastu | 3 | Light | 80 | 5 | 1 | 20.0% | 1.5% | 0.1 - 6.5 |

In Dhading, data were analysed from the 13-month collection period (September 2022-September 2023). Site 1 showed pool positivity of 10.3% (gravid) and 7.1% (light), with a corresponding infection prevalence (MLE) in *Cx. quinquefasciatus* of 0.5% for both trap types. Site 2 showed pool positivity of 16.1% (gravid) and 17.6% (light), with a corresponding infection prevalence (MLE) of 0.9% for both trap types. Infection estimates were similar between gravid and light traps at each site (Table 5).

In Jhapa and Kapilvastu, data were analysed over the 6-month period (March to August 2023). In Jhapa, pool positivity and infection prevalence (MLE) varied across sites and trap types, with prevalence (MLE) ranging from approximately 0.9% to 2.0% (Table 5). Higher prevalence estimates were generally observed in gravid traps compared with light traps at Site 3, while differences between trap types were smaller at Sites 1 and 2 (Table 5). In Kapilvastu, both pool positivity and infection prevalence (MLE) showed variation across sites and trap types, with infection prevalence ranging from approximately 0.3% to 1.5%. Prevalence estimates were broadly similar between gravid and light traps at most sites, with some variation observed at Site 3 (Table 5).

#### Temporal and climate analysis

Overall, monthly pool positivity and infection prevalence (MLE) in *Cx. quinquefasciatus* varied across the three study sites. Data are summarised in Table 5 for the 13-month periods in Dhading (2022-2023) and 6-month periods in Jhapa and Kapilvastu in 2023. To compare trends across the three study locations, data on the pool positivity and infection prevalence (MLE) for the 6-month period are presented graphically in Figs 5A–C. A similar graph for Dhading’s 13-month period is available in S1 File along with epidemiological week-wise data. Detailed data on the outputs of the PoolScreen analyses are provided S1 File.

**Figure 5.**
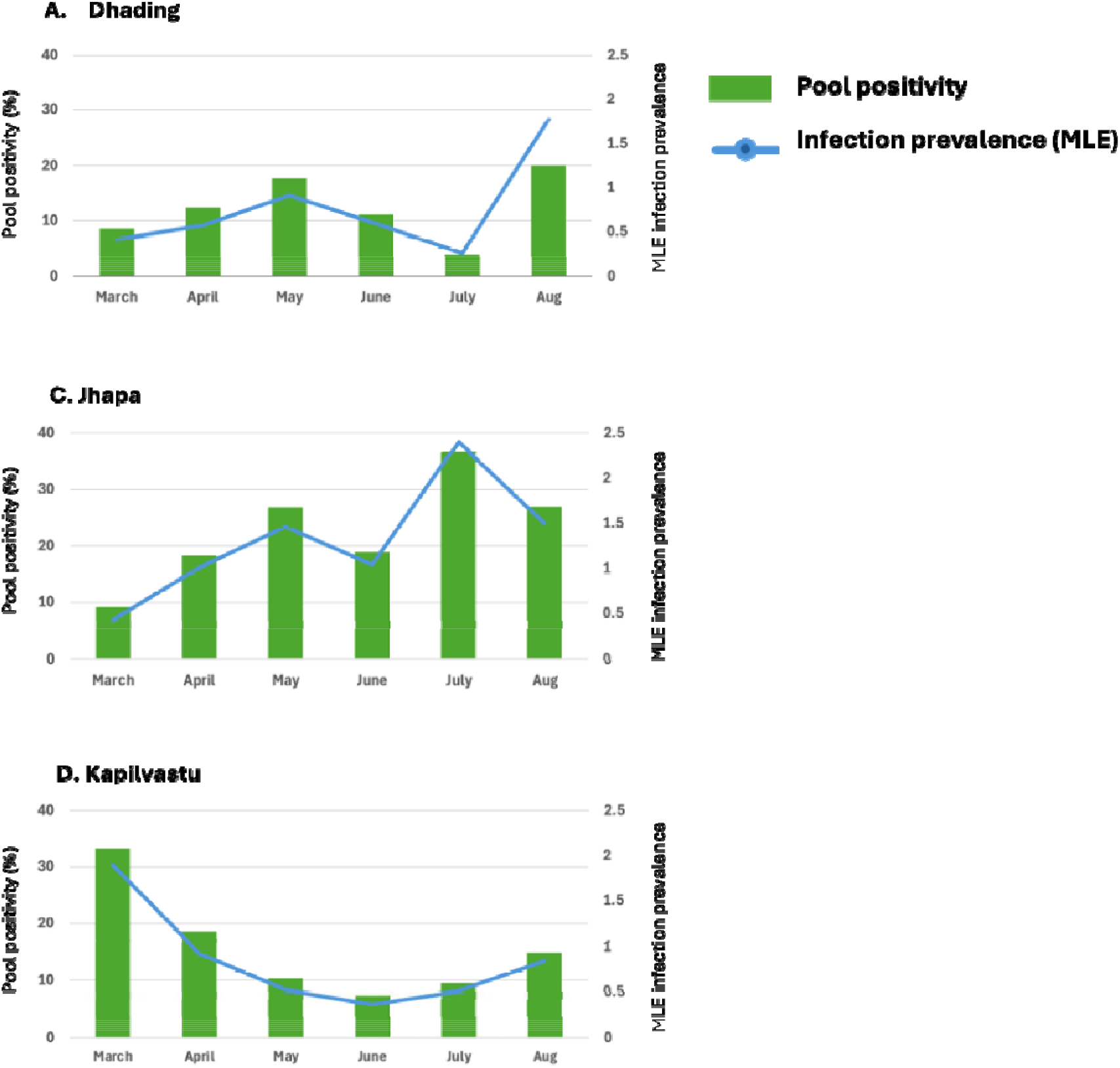
Monthly pool positivity and infection prevalence (MLE) in *Cx. quinquefasciatus*.

In Dhading, pool positivity increased from 8.5% in March to 20.0% in August 2023, showing a marked rise during the late monsoon period. Infection prevalence (MLE) followed a similar pattern, remaining low from March (0.4%) to July (0.3%), before increasing in August (1.8%) and peaking in August 2023 (1.8%) (Fig 5A). Pool positivity and infection prevalence (MLE) were significantly positively correlated (r = 0.968, p < 0.0001), indicating closely aligned temporal trends. In addition, overall positive associations were observed between MLE prevalence and precipitation and temperature, although these relationships were not statistically significant.

In Jhapa District, pool positivity increased fourfold between March (9.1%) and July (36.7%), while MLE prevalence increased steadily from March (0.4%) to June (1.0%), peaking in July (2.4%) before declining in August (1.5%) (Fig 5B). Pool positivity and Infection prevalence (MLE) were significantly positively correlated (r = 0.989, p < 0.0001). Positive associations were also observed between infection prevalence (MLE) and precipitation, temperature, and humidity; however, these relationships were not statistically significant.

In Kapilvastu District, pool positivity decreased from March (33.3%) to June (7.4%), with minor fluctuations in July and August. Infection prevalence (MLE) patterns were similar with decline from March (1.9%) to a small increase in August (0.8%) as shown in Fig 5C. Pool positivity and infection prevalence (MLE) were significantly positively correlated (r = 0.977, p < 0.001) (Fig 5C). Significant negative associations were observed between infection prevalence (MLE) and temperature (r = -0.855, p = 0.03), while associations with rainfall and humidity were not statistically significant.

## Discussion

This study demonstrates substantial spatial, seasonal, and site-specific heterogeneity in mosquito abundance and mosquito genus composition across three LF-endemic districts of Nepal. The dominance of *Culex* spp. across all districts underscores their potential role in LF transmission and supports their use as sentinel vectors for xenomonitoring, especially in low-endemic settings, with a focus on potential hotspots [6]. Understanding the relationship between infection rates in mosquitoes and the local at-risk human population is critically important, as mosquito infection rates may correlate with human infection pattern but can vary by vector species and ecological setting [39].

The WHO advocates for integrated vector control and surveillance [7,40], and as revealed in this study may help to confirm *W. bancrofti* circulating in communities. This is of particular concern in Dhading that stopped MDA in 2018 and may require additional investigation. Additional support to further integrate entomological surveillance within Nepal’s existing vector-borne disease control system would be beneficial [41]. This aligns with the WHO NTD roadmap’s integration component and could strengthen sustainability [1]. However, capacity and funding constraints remain critical challenges and represent an important gap, not only in Nepal but across many endemic countries globally [40].

Our analysis of species abundance and composition highlighted that, while *Culex* spp. were the dominant species, there was clear spatial variation in other species across locations. The relatively high proportion of *Anopheles* spp. in Kapilvastu suggests ecological conditions favourable for malaria vectors, while the presence of *Armigeres* spp. in Dhading may reflect the availability of organic-rich larval habitats unique to this Hill area. These district-level differences emphasise the influence of local environmental conditions in shaping mosquito compositions and are consistent with previous studies in Nepal, which demonstrate spatial and ecological heterogeneity in vector populations associated with the country’s diverse geography [20,21].

Gravid traps were significantly more efficient than light traps for capturing female *Cx. quinquefasciatus* across all study locations; however, some variation in trap efficiency was observed with higher rates in Jhapa. Because gravid females are epidemiologically most relevant for LF transmission, the better performance of gravid traps highlights their value for routine surveillance and post-MDA monitoring, also shown in studies elsewhere [6,8–10]. Light traps, although useful for general mosquito surveillance, were less selective for the LF *Culex* spp. vectors, and therefore may be less effective for targeted surveillance.

Our examination of *Cx. quinquefasciatus* within locations indicates potential household-level micro-hotspots of transmission. All locations showed significantly higher abundances in Site 2 than Site 1, suggesting that the housing characteristics, presence of livestock, and/or LF morbidity cases may be important factors driving localized transmission. While not all infected individuals develop clinical disease [42], the presence of LF morbidity cases may help identify higher-risk areas as shown elsewhere [43,44]. A micro-stratification approach has been implemented for malaria control in Nepal incorporating disease burden, ecology/vector transmission, and population movement to identify risk areas and guide interventions [45]. A similar approach for LF, applied at a finer geographical scale, could improve the targeted placement of surveillance and control tools, such as gravid traps in higher-risk sites.

Temporal and seasonal analysis indicates that LF transmission risk is likely highest during the pre-monsoon and monsoon periods, when vector abundance is persistent. Peak episodes were more pronounced n Dhading than in Jhapa or Kapilvastu, likely reflecting ecological differences between Nepal’s Hill and Terai ecological regions [19]. The alignment between seasonal mosquito abundance and persistent monthly LF pool positivity underscores the importance of timing surveillance and control interventions to periods of highest risk and highlights the need to improve our understanding of climate drivers.

Although the climate analysis was exploratory and limited to monthly data over a relatively short period, rainfall, temperature, and humidity appeared to influence both mosquito abundance and infection rates. This warrants more detailed analysis using longer-term datasets and finer temporal resolution (e.g. weekly or daily data). Vector borne disease surveillance will become increasingly important as climate change alters vector distributions in Nepal, with several vector-borne diseases already being reported at higher altitudes [19,24]. Closer collaboration between vector-borne disease programmes and climate organisations could further strengthen predictive surveillance and help identify areas at greatest future risk.

Overall, the detection of LF-positive mosquito pools across all districts is of concern and indicates ongoing *W. bancrofti* transmission, with Jhapa emerging as a higher-risk focus. Gravid traps generally yielded slightly higher pool positivity and infection prevalence (MLE) than light traps, particularly in Jhapa and Kapilvastu. Temporal analysis also revealed that while infection prevalence infections were relatively low overall, there were periodic peaks in selected months and epidemiological weeks. These findings highlight the continued importance of entomological surveillance, particularly the use of gravid traps during high-risk transmission periods in micro-stratified high-risk locations, to guide targeted vector control and support LF elimination efforts in Nepal and elsewhere [1].

The main limitations of this study include its exploratory design, coverage of only one site per district, differences in sampling duration between study locations, and restriction to a single season/year due to funding constraints. In addition, the climate analysis was exploratory and based on a relatively short period of monthly data, limiting the ability to fully characterise climate–vector relationships.

## Conclusion

This study provides new insights into the spatial and temporal heterogeneity of mosquito populations, particularly *Cx. quinquefasciatus*, and their relationship with environmental factors in LF-endemic areas of Nepal. It highlights the utility and effectiveness of gravid trap– based xenomonitoring as a tool for assessing vector abundance and *W. bancrofti* infection. These findings will help inform national LF elimination and vector-borne disease surveillance programmes and support targeted vector control strategies by optimising the timing and targeting of interventions.

## Supporting information

(see S1 checklist).

## Data Availability

The data used are available in the manuscript and supplementary files. Additional data are available from the corresponding author upon reasonable request.

## Supporting information

S1 Checklist. STROBE checklist

S1 File. Summary information on methods and selected results.

## Authors contributions

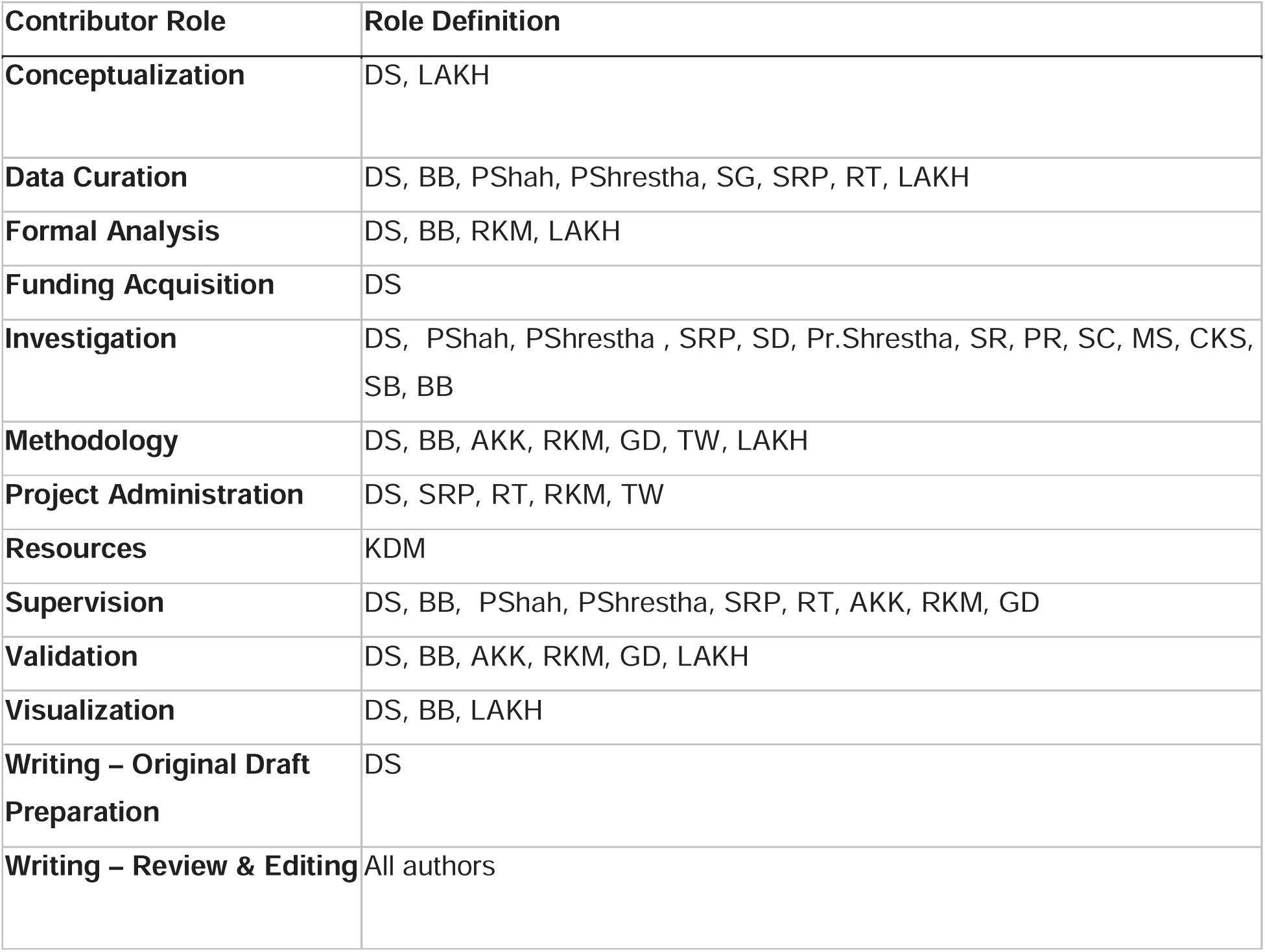

## Acknowledgements

We are grateful to the householders who permitted access to their premises to conduct this study. We sincerely thank the local field workers who managed the daily mosquito collections, and to the local community for facilitating site access.

We thank Dr Themba Mzilahowa from the Malaria Alert Centre (MAC), of the Kamuzu University of Health Sciences (KUHeS), Blantyre, Malawi for providing *Wuchereria bancrofti* sample concentration, DNA extraction, and the PCR protocol used to determine infection in *Culex* mosquitoes collected during the study period.

The study was supported by a grant from GSK awarded to DS and forms part of the project entitled ‘GSK-Microsoft Premonition to Accelerate Global Lymphatic Filariasis Elimination: a pilot study in Nepal’. We acknowledge all the members of the Microsoft team for productive discussions and for guidance on mosquito infection calculations.

The authors used ChatGPT (GPT-5.5, OpenAI) to improve readability of selected written text and for checking scripts in R Studio. The authors reviewed and edited the content as needed and take full responsibility for the content of the published manuscript.

