## Supplementary material for "Longitudinal entomological study using molecular xenomonitoring of *Wuchereria bancrofti* in *Culex quinquefasciatus* in three Nepalese districts: Implications for lymphatic filariasis transmission assessment and elimination": S1 file

**Supplementary information**

**Methods related data**

**S1. Table. Summary of site characteristics and trap type within each of the three study locations**

| **Site No.** | **Trap type** | **Dhading** | **Jhapa** | **Kapilvastu** |
| --- | --- | --- | --- | --- |
| **Site 1** | **Light trap** | Outdoor, near the animal shed | Indoor, inside the house where field officers were staying | Indoor, in the passage of lab room |
| **Site 1** | **Gravid trap** | Outdoor, near stagnant water source from hand pump | Outdoor, outside the house in the garden area | Outdoor, in the garden of primary health centre near sewage tank |
| **Site 2** | **Light trap** | Indoor, inside the LF patient’s house | Indoor, inside the LF patient’s house | Indoor, inside the house near to patient’s house |
| **Site 2** | **Gravid trap** | Outdoor, outside the house near stagnant water | Outdoor, outside the house in the garden area | Outdoor, behind the house near the pond |
| **Site 3** | **Light trap** | NA | Indoor, inside the LF patient’s house | Indoor, inside the house near to LF patient’s house |
| **Site 3** | **Gravid trap** | NA | Outdoor, outside the house near stagnant water | Outdoor, in the garden area |

**Protocol for the detection of *Wuchereria bancrofti* in female *Culex quinquefasciatus***

The PCR amplification was performed using *W. bancrofti* specific primers LDR1 and LDR2 real time primers for real time detection. Real-time PCR reactions were performed using 12.5 µl of TaqMan master mix (Applied Biosystems, USA) along with 450 nmol/l of each primer and 125 nmol/l probe in a final volume of 25 µl in 96-well MicroAmp optical plates (Applied Biosystems, USA). One micro litre of the extracted DNA (by Qiagen) was used as template in real-time PCR. Thermal cycling parameters used were 50°C for 2 min, 95°C for 10 min followed by 40 cycles of 95°C for 15 sec and 60°C for 1 min (15). Positive and negative controls were also run in each run. Thermal cycling and data analysis were done with Thermal cycler (Biorad, Germany) using CFX Maestro software. Cycle threshold (Ct) values of samples ranging from 1–38 was considered positive, and samples that failed to reach the fluorescence threshold beyond 38 were considered indeterminate and repeated to confirm the negativity or positivity of those samples as described by Rao et al., 2006 [30].

**S2. Table****. Sequence of primers and probe**

| **Primer / Probes** | **Sequence** |
| --- | --- |
| LDR1 | 5’ATTTTGATCATCTGGGAACGTTAATA-3’ |
| LDR2 | 5’CGACTGTCTAATCCATTCAGAGTGA-3’ |
| Probe | 6 FAM-ATCTGCCCATAG AAATAACT ACGGTGGATCTG-TAMRA |

**Objective 1 related data. Characterize the abundance, species composition of mosquitoes**

**S3. Table. Species of mosquitoes trapped by different devices in three districts**

| **Districts** | **Species of mosquitoes trapped by light trap** | **Species of mosquitoes trapped by Gravid trap** |
| --- | --- | --- |
| **Dhading** | *Culex quinquefasciatus*  *Culex fuscocephala*  *Culex epidesmus*  *Culex gelidus*  *Culex whitemori*  *Culex hutchinsoni*  *Culex vishnui*  *Culex fuscanus*  *Culex epidesmus*  *Culex jacsoni*  *Culex vagans*  *Culex bitaeniorhynchus*  *Culex tritaeniorhynchus*  *Culex mimeticus*  *Culex mimulus*  *Culex pseudovishnui*  *Culex Vishnui*  *Culex (lutzia)* spp  *Culex* spp.  *Anopheles barbirostris*  *Anopheles pallidus*  *Anopheles sinensis*  *Anopheles dravidicus.*  *Anopheles maculatus complex*  *Anopheles subpictus*  *Anopheles vagus*  *Anopheles pseudowillmori*  *Anopheles splendidus*  *Aedes.albopictus*  *Aedes subalbopictus*  *Aedes lineatopennis*  *Aedes pallidostriatus*  *Aedes pulchriventer*  *Aedes pipersalatus*  *Aedes vexans*  *Aedes walbus*  *Aedes imitator*  *Aedes aegypti*  *Armigeres* spp.  *Armigeres subalbatus*  *Armigeres kuchingensis*  *Malaya* spp.  *Mansonia* spp*.* | *Culex quinquefasciatus*  *Culex fuscocephala*  *Culex gelidus*  *Culex hutchinsoni*  *Culex epidesmus*  *Culex fuscanus*  *Culex bitaeniorhynchus*  *Culex Vagans*  *Culex (lutzia)* spp  *Anopheles splendidus*  *Anopheles dravidicus*  *Anopheles mac dravidicus*  *Anopheles sinensis*  *Anopheles* spp*.*  *Anopheles vagus*  *Anopheles annularis*  *Aedes vittatus*  *Aedes albopictus*  *Aedes subalbopictus*  *Aedes* spp.  *Aedes pallidostriatus*  *Aedes pipersalatus*  *Armigeres* spp*.*  *Armigeres kuchingensis*  *Armigeres subalbatus* |
| **Jhapa** | *Culex quinquefasciatus*  *Culex tritaeniorhynchus*  *Culex gelidus*  *Culex fuscocephala*  *Culex* spp*.*  *Culex whitemorei*  *Culex vishnui*  *Mansonia* spp.  *Anopheles* spp.  *Aedes* spp*.*  *Armigeres* spp. | *Culex quinquefasciatus*  *Culex tritaeniorhynchus*  *Culex fuscocephala*  *Culex bitaeniorhynchus*  *Culex* spp.  *Culex gelidus*  *Culex (lutzia)* spp  *Culex whitemorei*  *Culex vishnui*  *Culex pseudovishnui*  *Armigeres* spp*.*  *Anopheles* spp*.*  *Aedes* spp.  *Aedes albopictus*  *Mansonia* spp.  *Mansonia annularis*  *Mansonia uniformis*  *Toxorhynchites* spp*.* |
| **Kapilvastu** | *Culex quinquefasciatus*  *Culex fuscocephala*  *Culex* spp*.*  *Culex pseudovishnui*  *Culex whitemorei*  *Culex vishnui*  *Anopheles aconitus*  *Anopheles barbirostris*  *Anopheles pallidus*  *Anopheles vagus*  *Anopheles sinensis*  *Anopheles subpictus*  *Anopheles culicifacies*  *Anopheles annularis*  *Anopheles fluviatilis*  *Armigeres* spp.  *Aedes* spp.  *Mansonia* spp. | *Culex quinquefasciatus*  *Culex fuscocephala*  *Culex* spp.  *Culex pseudovishnui*  *Culex whitemorei*  *Culex vishnui*  *Anopheles subpictus*  *Anopheles culicifacies*  *Anopheles annularis*  *Anopheles fluviatilis*  *Armigeres* spp*.*  *Aedes* spp*.*  *Mansonia* spp*.* |

**Objective 2 related data. Compare *Cx. quinquefasciatus* abundance and gravidity by trap type.**

**S4. Table****: Dhading District – summary of female and gravid *Culex quinquefasciatus* numbers by trap type and placement**

| **Site No.** | **District** | **Type of device used and placement** | **Site** | **Number of total mosquitoes collected** | **No. of female *Culex quinquefasciatus*** | **No. of gravid female *Culex quinquefasciatus*** |
| --- | --- | --- | --- | --- | --- | --- |
| 1. | Dhading | Gravid trap (GV1)  Outdoor | 1 | 5,387 | 1,605 | 1,355 |
| 2. | Dhading | Gravid trap (GV2)  Outdoor | 2 | 4,199 | 2,289 | 1,764 |
| 3. | Dhading | Light trap (LT1)  Indoor | 1 | 4,605 | 630 | 104 |
| 4. | Dhading | Light trap (LT2) indoor | 2 | 12,805 | 2,641 | 581 |
| **Total** | | | | **26,996** | **7,165** | **3,804** |

Note: Nightly collections between 18^th^ September 2022 and 17^th^ September 2023

**Statistical comparison by trap type**

| **Trap type** | **No. of female *Culex quinquefasciatus*** | **No. of gravid female *Culex quinquefasciatus*** | **Percentage (%) of gravid *Culex quinquefasciatus*** |
| --- | --- | --- | --- |
| Gravid | 3894 | 3119 | 80.1% |
| Light | 3271 | 685 | 20.9% |
| **Total** | **7165** | **3894** |  |

Mann-Whitney U = 34101, p<0.001

**S5.Table****: Jhapa District – summary of female and gravid *Culex quinquefasciatus* numbers by trap type and placement**

| **Site No.** | **District** | **Type of device used and placement** | **Site** | **Number of total mosquitoes collected** | **No. of female *Culex quinquefasciatus*** | **No. of gravid female *Culex quinquefasciatus*** |
| --- | --- | --- | --- | --- | --- | --- |
| 1. | Jhapa | Gravid trap (GV1)  Outdoor | 1 | 3,316 | 1,603 | 633 |
| 2. | Jhapa | Gravid trap (GV2)  Outdoor | 2 | 3,202 | 1,583 | 635 |
| 3. | Jhapa | Gravid trap (GV3)  Outdoor | 3 | 1,458 | 890 | 803 |
| 4. | Jhapa | Light trap (LT1)  Indoor | 1 | 3,613 | 222 | 55 |
| 5. | Jhapa | Light trap (LT2)  Indoor | 2 | 3,229 | 781 | 89 |
| 6. | Jhapa | Light trap (LT3)  Indoor | 3 | 2,203 | 129 | 19 |
| **Total** | | | | **17,021** | **5,208** | **2,234** |

**Note:** Nightly collections in site 1 and site 2 between 26th March 2023 and 31st August 2023. Nightly collections in site 3 between 25^th^ July and 31st August 2023.

**S6.Table: Statistical comparison by trap type**

| **Trap type** | **No. of female *Culex quinquefasciatus*** | **No. of gravid female *Culex quinquefasciatus*** | **Percentage (%) of gravid *Culex quinquefasciatus*** |
| --- | --- | --- | --- |
| Gravid | 4076 | 2071 | 50.8% |
| Light | 1132 | 163 | 14.4% |
| **Total** | **5208** | **2234** |  |

Mann-Whitney U = 6541, p<0.001

**S7 Table****: Kapilvastu District – summary of female and gravid *Culex quinquefasciatus* numbers by trap type and placement**

| **Site No.** | **District** | **Trap type and placement** | **Site** | **Number of total mosquitoes collected** | **No. of female *Culex quinquefasciatus*** | **No. of gravid female *Culex quinquefasciatus*** |
| --- | --- | --- | --- | --- | --- | --- |
| 1. | Kapilvastu | Gravid trap (GV1)  Outdoor | 1 | 1,854 | 470 | 199 |
| 2. | Kapilvastu | Gravid trap (GV2)  Outdoor | 2 | 4,768 | 1,686 | 779 |
| 3. | Kapilvastu | Gravid trap (GV3)  Outdoor | 3 | 781 | 173 | 138 |
| 4. | Kapilvastu | Light trap (LT1)  Indoor | 1 | 2,060 | 323 | 76 |
| 5. | Kapilvastu | Light trap (LT2)  Indoor | 2 | 5,808 | 939 | 346 |
| 6. | Kapilvastu | Light trap (LT3) Indoor | 3 | 1,595 | 80 | 37 |
| **Total** | | | | **16,866** | **3,671** | **1,575** |

**Note:** Nightly collections in site 1 and site 2 between 26th March 2023 and 31st August 2023. Nightly collections in site 3 between 6^th^ August 2023 and 31st August 2023.

**S8. Table: Statistical comparison by trap type**

| **Trap type** | **No. of female *Culex quinquefasciatus*** | **No. of gravid female *Culex quinquefasciatus*** | **Percentage (%) of gravid *Culex quinquefasciatus*** |
| --- | --- | --- | --- |
| Gravid | 2329 | 1116 | 47.9% |
| Light | 1342 | 459 | 34.2% |
| **Total** | **3671** | **1575** |  |

Mann-Whitney U = 13604, p<0.001

**Objective 3 related data**

**S9. Table: Dhading District site and trap specific *Cx. quinquefasciatus* distributions**

**Full 13 months collections**

| **District and**  **Trap type** | **Other species** | | | ***Cx. quinquefasciatus*** | | |  |
| --- | --- | --- | --- | --- | --- | --- | --- |
| **Dhading** | **Total** | **Male** | **Female** | **Total** | **Male** | **Female** | **Overall total** |
| GV1  Gravid - Site 1 | 3576 | 511 | 5387 | 1811 | 206 | 1605 | 5387 |
| GV2  Gravid - Site 2 | 1661 | 192 | 4199 | 2538 | 249 | 2289 | 4199 |
| LT1  Light - Site 1 | 3848 | 776 | 4605 | 757 | 127 | 630 | 4605 |
| LT2  Light - Sight 2 | 8693 | 7589 | 12805 | 4112 | 1471 | 2641 | 12805 |
| **Grand Total** | **17778** | **9068** | **26996** | **9218** | **2053** | **7165** | **26996** |

**S10. Table: 6 months collections**

| **District and**  **Trap type** | **Other species** | | | ***Cx. quinquefasciatus*** | | |  |
| --- | --- | --- | --- | --- | --- | --- | --- |
| **Dhading** | **Total** | **Male** | **Female** | **Total** | **Male** | **Female** | **Overall total** |
| GV1  Gravid - Site 1 | 2008 | 379 | 1629 | 1398 | 158 | 1240 | 3406 |
| GV2  Gravid - Site 2 | 636 | 181 | 455 | 2045 | 217 | 1828 | 2681 |
| LT1  Light - Site 1 | 1339 | 600 | 739 | 673 | 97 | 576 | 2012 |
| LT2  Light - Sight 2 | 8169 | 7421 | 635 | 3570 | 1250 | 2320 | 11739 |
| **Grand Total** | **12152** | **8581** | **3458** | **7686** | **1722** | **5964** | **19838** |

**S11. Table: Jhapa District site and trap specific *Cx. quinquefasciatus* distributions**

**6 months collections of two main trap sites**

| **District and**  **Trap type** | **Other species** | | | ***Cx. quinquefasciatus*** | | |  |
| --- | --- | --- | --- | --- | --- | --- | --- |
| **Dhading** | **Total** | **Male** | **Female** | **Total** | **Male** | **Female** | **Overall total** |
| GV1  Gravid - Site 1 | 1713 | 855 | 865 | 1603 | 0 | 1603 | 3316 |
| GV2  Gravid - Site 2 | 1619 | 899 | 716 | 1583 | 0 | 1583 | 3202 |
| LT1  Light - Site 1 | 3391 | 571 | 2809 | 222 | 0 | 222 | 3613 |
| LT2  Light - Sight 2 | 2448 | 731 | 1719 | 781 | 0 | 781 | 3229 |
| **Grand Total** | **9171** | **3056** | **6109** | **4189** | **0** | **4189** | **13360** |

**S12. Table: Kapilvastu District site and trap specific *Cx. quinquefasciatus* distributions**

**6 months collections of two main trap sites**

| **District and**  **Trap type** | **Other species** | | | ***Cx. quinquefasciatus*** | | |  |
| --- | --- | --- | --- | --- | --- | --- | --- |
| **Dhading** | **Total** | **Male** | **Female** | **Total** | **Male** | **Female** | **Overall total** |
| GV1  Gravid - Site 1 | 1384 | 969 | 415 | 470 | 0 | 470 | 1854 |
| GV2  Gravid - Site 2 | 3082 | 2380 | 702 | 1686 | 0 | 1686 | 4768 |
| LT1  Light - Site 1 | 1737 | 775 | 962 | 323 | 0 | 323 | 2060 |
| LT2  Light - Sight 2 | 4869 | 2059 | 2810 | 939 | 0 | 939 | 5808 |
| **Grand Total** | **11072** | **6183** | **4889** | **3418** | **0** | **3418** | **14490** |

**Objective 4 related data. Temporal and seasonal patterns of *Culex quinquefasciatus* abundance and associated climatic factors**

**S13. Table. Summary of monthly total and daily range of female *Culex quinquefasciatus* by trap type and associated monthly climate measures**

| **District** | **Gravid trap**  **Total number**  **(Daily range)** | **Light trap**  **Total number**  **(Daily range)** | **Monthly Total number** | **Precip.**  **total (mm)** | **Temp.**  **average (°C)** | **Humidityaverage (%)** |
| --- | --- | --- | --- | --- | --- | --- |
| **Dhading** | | | | | | |
| Sept-22 | 24 (1-6) | 148 (3-50) | 172 | 92.7 | 24.6 | 68.8 |
| Oct-22 | 274 (2-42) | 105 (2-15) | 379 | 133.1 | 21.2 | 65.1 |
| Nov-22 | 217 (1-48) | 44 (1-5) | 261 | 0.7 | 19.7 | 63.7 |
| Dec-22 | 107 (0-10) | 6 (0-4) | 113 | 8.0 | 15.0 | 78.5 |
| Jan-23 | 10 (0-2) | 0 (0-0) | 10 | 7.6 | 13.1 | 78.1 |
| Feb-23 | 70 (0-8) | 24 (0-4) | 94 | 8.5 | 15.5 | 72.4 |
| Mar-23 | 661 (1-71) | 374 (0-44) | 1035 | 53.2 | 18.0 | 67.9 |
| Apr-23 | 975 (1-75) | 1351 (6-161) | 2326 | 30.8 | 21.2 | 59.4 |
| May-23 | 464 (2-50) | 763 (1-65) | 1227 | 126.7 | 22.2 | 77.3 |
| Jun-23 | 488 (1-53) | 257 (2-21) | 745 | 201.1 | 26.2 | 73.9 |
| Jul-23 | 278 (2-36) | 82 (1-9) | 360 | 348.8 | 26.8 | 83.8 |
| Aug-23 | 202 (1-22) | 69 (1-9) | 271 | 408.5 | 25.5 | 86.8 |
| Sep-23 | 124 (3-31) | 48 (1-8) | 172 | 166.0 | 26.2 | 83.6 |

| **Jhapa** | | | | | | |
| --- | --- | --- | --- | --- | --- | --- |
| Mar-23 | 238 (30-73) | 7 (0-7) | 245 | 13.0 | 28.5 | 31.2 |
| Apr-23 | 661 (3-68) | 74 (1-7) | 735 | 36.6 | 31.5 | 32.9 |
| May-23 | 916 (2-123) | 165 (1-73) | 1081 | 111.3 | 32.2 | 31.6 |
| Jun-23 | 395 (1-59) | 427 (1-159) | 822 | 228.1 | 32.6 | 42.7 |
| Jul-23 | 615 (1-75) | 225 (1-79) | 840 | 706.2 | 32.4 | 47.0 |
| Aug-23 | 1251 (5-141) | 234 (1-34) | 1485 | 576.8 | 32.1 | 47.2 |

| **Kapilvastu** | | | | | | |
| --- | --- | --- | --- | --- | --- | --- |
| Mar-23 | 58 (7-20) | 0 | 58 | 6.9 | 28.1 | 34.4 |
| Apr-23 | 624 (5-48) | 19 (8-11) | 643 | 26.0 | 30.2 | 33.1 |
| May-23 | 395 (4-39) | 316 (3-22) | 711 | 76.4 | 30.9 | 39.2 |
| Jun-23 | 389 (4-32) | 259 (1-20) | 648 | 202.9 | 34.1 | 40.5 |
| Jul-23 | 379 (1-33) | 408 (2-53) | 787 | 367.0 | 31.3 | 60.2 |
| Aug-23 | 484 (3-46) | 340 (3-34) | 58 | 6.9 | 28.1 | 34.4 |

**S14.Table: Summary of Epi-week number of female *Culex quinquefasciatus***

| **Year, Month, Epi-Week** | **Dhading** | **Jhapa** | **Kapilvastu** |
| --- | --- | --- | --- |
| 2022SepEPI 38 | 49 |  |  |
| 2022SepEPI 39 | 123 |  |  |
| 2022OctEPI 40 | 15 |  |  |
| 2022OctEPI 41 | 89 |  |  |
| 2022OctEPI 42 | 191 |  |  |
| 2022OctEPI 43 | 84 |  |  |
| 2022NovEPI 45 | 52 |  |  |
| 2022NovEPI 46 | 134 |  |  |
| 2022NovEPI 47 | 64 |  |  |
| 2022NovEPI 48 | 11 |  |  |
| 2022DecEPI 48 | 7 |  |  |
| 2022DecEPI 49 | 44 |  |  |
| 2022DecEPI 50 | 46 |  |  |
| 2022DecEPI 51 | 8 |  |  |
| 2022DecEPI 52 | 8 |  |  |
| 2023JanEPI 1 | 5 |  |  |
| 2023JanEPI 2 | 1 |  |  |
| 2023JanEPI 3 | 1 |  |  |
| 2023JanEPI 4 | 2 |  |  |
| 2023JanEPI 5 | 1 |  |  |
| 2023FebEPI 5 | 2 |  |  |
| 2023FebEPI 6 | 15 |  |  |
| 2023FebEPI 7 | 21 |  |  |
| 2023FebEPI 8 | 31 |  |  |
| 2023FebEPI 9 | 25 |  |  |
| 2023MarEPI 9 | 3 |  |  |
| 2023MarEPI 10 | 61 |  |  |
| 2023MarEPI 11 | 330 |  |  |
| 2023MarEPI 12 | 257 |  |  |
| 2023MarEPI 13 | 384 | 245 | 58 |
| 2023AprilEPI 14 | 292 | 171 | 110 |
| 2023AprilEPI 15 | 588 | 129 | 170 |
| 2023AprilEPI 16 | 829 | 153 | 118 |
| 2023AprilEPI 17 | 540 | 210 | 211 |
| 2023AprilEPI 18 | 77 | 72 | 34 |
| 2023MayEPI 18 | 338 | 378 | 126 |
| 2023MayEPI 19 | 124 | 353 | 121 |
| 2023MayEPI 20 | 276 | 115 | 192 |
| 2023MayEPI 21 | 386 | 160 | 155 |
| 2023MayEPI 22 | 103 | 75 | 117 |
| 2023JuneEPI 22 | 37 | 63 | 68 |
| 2023JuneEPI 23 | 179 | 141 | 217 |
| 2023JuneEPI 24 | 207 | 228 | 147 |
| 2023JuneEPI 25 | 174 | 102 | 107 |
| 2023JuneEPI 26 | 148 | 288 | 109 |
| 2023JulyEPI 27 | 149 | 146 | 82 |
| 2023JulyEPI 28 | 83 | 236 | 270 |
| 2023JulyEPI 29 | 47 | 291 | 214 |
| 2023JulyEPI 30 | 69 | 141 | 179 |
| 2023JulyEPI 31 | 12 | 26 | 42 |
| 2023AugEPI 31 | 37 | 121 | 41 |
| 2023AugEPI 32 | 45 | 291 | 210 |
| 2023AugEPI 33 | 29 | 392 | 154 |
| 2023AugEPI 34 | 84 | 485 | 287 |
| 2023AugEPI 35 | 76 | 196 | 132 |
| 2023SepEPI 35 | 20 |  |  |
| 2023SepEPI 36 | 79 |  |  |
| 2023SepEPI 37 | 62 |  |  |
| 2023SepEPI 38 | 11 |  |  |

**Correlations between daily mean abundance and climate factors**

**Dhading – 13 months**

| **Correlations** | | | | | | | |
| --- | --- | --- | --- | --- | --- | --- | --- |
|  | | Gravid Trap Daily mean | Light Trap Daily mean | Total Daily mean | Precip.  (mm) total | Temp  Mean | Humidity (%) mean |
| Gravid Trap Daily mean | Pearson Correlation | 1 | .819^**^ | .928^**^ | -.047 | .199 | -.550 |
|  | Sig. (2-tailed) |  | <.001 | <.001 | .879 | .514 | .052 |
|  | N | 13 | 13 | 13 | 13 | 13 | 13 |
| Light Trap Daily mean | Pearson Correlation | .819^**^ | 1 | .974^**^ | -.179 | .129 | -.517 |
|  | Sig. (2-tailed) | <.001 |  | <.001 | .559 | .676 | .070 |
|  | N | 13 | 13 | 13 | 13 | 13 | 13 |
| Total Daily mean | Pearson Correlation | .928^**^ | .974^**^ | 1 | -.120 | .169 | -.541 |
|  | Sig. (2-tailed) | <.001 | <.001 |  | .696 | .581 | .056 |
|  | N | 13 | 13 | 13 | 13 | 13 | 13 |
| **. Correlation is significant at the 0.01 level (2-tailed). | | | | | | | |
| *. Correlation is significant at the 0.05 level (2-tailed). | | | | | | | |

**Pearson Correlations** ██ Highly Positive ██ Positive ██ No Linear Correlation ██ Negative: ██ Highly Negative

**Dhading – 6 months**

|  | **Correlations** | | | | | | | |
| --- | --- | --- | --- | --- | --- | --- | --- | --- |
|  | | | Gravid Trap Daily mean | Light Trap Daily mean | Total Daily mean | Precip.  (mm) total | Temp  Mean | Humidity (%) mean |
| Gravid Trap Daily mean | | Pearson Correlation | 1 | .855^*^ | .932^**^ | -.889^*^ | -.653 | -.994^**^ |
|  |  | Sig. (2-tailed) |  | .030 | .007 | .018 | .159 | <.001 |
|  |  | N | 6 | 6 | 6 | 6 | 6 | 6 |
| Light Trap Daily mean | | Pearson Correlation | .855^*^ | 1 | .985^**^ | -.759 | -.519 | -.813^*^ |
|  |  | Sig. (2-tailed) | .030 |  | <.001 | .080 | .291 | .049 |
|  |  | N | 6 | 6 | 6 | 6 | 6 | 6 |
| Total Daily mean | | Pearson Correlation | .932^**^ | .985^**^ | 1 | -.823^*^ | -.573 | -.900^*^ |
|  |  | Sig. (2-tailed) | .007 | <.001 |  | .044 | .235 | .015 |
|  |  | N | 6 | 6 | 6 | 6 | 6 | 6 |

**. Correlation is significant at the 0.01 level (2-tailed).

*. Correlation is significant at the 0.05 level (2-tailed).

**Pearson Correlations** ██ Highly Positive ██ Positive ██ No Linear Correlation ██ Negative: ██ Highly Negative

Jhapa – 6 months

| **Correlations** | | | | | | | |
| --- | --- | --- | --- | --- | --- | --- | --- |
|  | | Gravid Trap Daily mean | Light Trap Daily mean | Total Daily mean | Precip.  (mm) total | Temp  Mean | Humidity (%) mean |
| Gravid Trap Daily mean | Pearson Correlation | 1 | -.436 | .905^*^ | -.062 | -.638 | -.239 |
|  | Sig. (2-tailed) |  | .387 | .013 | .908 | .173 | .649 |
|  | N | 6 | 6 | 6 | 6 | 6 | 6 |
| Light Trap Daily mean | Pearson Correlation | -.436 | 1 | -.065 | .298 | .344 | .566 |
|  | Sig. (2-tailed) | .387 |  | .902 | .566 | .504 | .242 |
|  | N | 6 | 6 | 6 | 6 | 6 | 6 |
| Total Daily mean | Pearson Correlation | .905^*^ | -.065 | 1 | .157 | -.394 | .084 |
|  | Sig. (2-tailed) | .013 | .902 |  | .766 | .439 | .874 |
|  | N | 6 | 6 | 6 | 6 | 6 | 6 |

**. Correlation is significant at the 0.01 level (2-tailed).

*. Correlation is significant at the 0.05 level (2-tailed)

**Pearson Correlations** ██ Highly Positive ██ Positive ██ No Linear Correlation ██ Negative: ██ Highly Negative

Kapilvastu – 6 months

|  | | Gravid Trap Daily mean | Light Trap Daily mean | Total Daily mean | Precip.  (mm) total | Temp  Mean | Humidity (%) mean |
| --- | --- | --- | --- | --- | --- | --- | --- |
| Gravid Trap Daily mean | Pearson Correlation | 1 | .062 | .188 | -.293 | -.200 | -.231 |
|  | Sig. (2-tailed) |  | .907 | .721 | .572 | .704 | .660 |
|  | N | 6 | 6 | 6 | 6 | 6 | 6 |
| Light Trap Daily mean | Pearson Correlation | .062 | 1 | .964^**^ | .728 | .536 | .655 |
|  | Sig. (2-tailed) | .907 |  | .002 | .101 | .273 | .158 |
|  | N | 6 | 6 | 6 | 6 | 6 | 6 |
| Total Daily mean | Pearson Correlation | .188 | .964^**^ | 1 | .756 | .460 | .728 |
|  | Sig. (2-tailed) | .721 | .002 |  | .082 | .358 | .101 |
|  | N | 6 | 6 | 6 | 6 | 6 | 6 |

**. Correlation is significant at the 0.01 level (2-tailed).

*. Correlation is significant at the 0.05 level (2-tailed).

**Pearson Correlations** ██ Highly Positive ██ Positive ██ No Linear Correlation ██ Negative: ██ Highly Negative

**Objective 5 related data. Infection data related to *Culex quinquefasciatus***

**S15-17. .Site and trap-specific analysis**

**S15. Table. Dhading**

| **Site** | **Trap** | **PrevMLE** | **CILow** | **CIHigh** | **NumberOfPools** | **NumberPositive** |
| --- | --- | --- | --- | --- | --- | --- |
| 1 | Gravid trap | 0.00526945 | 0.00241072 | 0.00979154 | 78 | 8 |
| 1 | Light trap | 0.00483979 | 0.00120568 | 0.01250364 | 42 | 3 |
| 2 | Gravid trap | 0.00864475 | 0.00524651 | 0.01325642 | 112 | 18 |
| 2 | Light trap | 0.0091189 | 0.00582581 | 0.01345877 | 125 | 22 |

**S16. Table. Jhapa**

| **Site** | **Trap** | **PrevMLE** | **CILow** | **CIHigh** | **NumberOfPools** | **NumberPositive** |
| --- | --- | --- | --- | --- | --- | --- |
| 1 | Gravid trap | 0.01218009 | 0.00727571 | 0.01889519 | 74 | 17 |
| 1 | Light trap | 0.01475795 | 0.00368565 | 0.03792497 | 17 | 3 |
| 2 | Gravid trap | 0.01521056 | 0.00330000 | 0.02350000 | 71 | 20 |
| 2 | Light trap | 0.01333928 | 0.00641761 | 0.02400055 | 42 | 9 |
| 3 | Gravid trap | 0.01977116 | 0.01113681 | 0.03201534 | 38 | 14 |
| 3 | Light trap | 0.00857985 | 0.00049065 | 0.03747078 | 8 | 1 |

**S17. Table. Kapilvastu**

| **Site** | **Trap** | **PrevMLE** | **CILow** | **CIHigh** | **NumberOfPools** | **NumberPositive** |
| --- | --- | --- | --- | --- | --- | --- |
| 1 | Gravid trap | 0.00674197 | 0.00168016 | 0.01740514 | 25 | 3 |
| 1 | Light trap | 0.00317025 | 0.00018114 | 0.01388862 | 19 | 1 |
| 2 | Gravid trap | 0.00777516 | 0.0041681 | 0.01301869 | 77 | 12 |
| 2 | Light trap | 0.00448626 | 0.00139443 | 0.01039778 | 45 | 4 |
| 3 | Gravid trap | 0.00618326 | 0.00035356 | 0.02701678 | 7 | 1 |
| 3 | Light trap | 0.01487598 | 0.00085013 | 0.06510472 | 5 | 1 |

**Temporal analyses**

**S18. Table. Dhading - Outputs from the PoolScreen analysis in R studio and pool positivity summaries**

|  |  | **Poolscreen analysis –infection prevalence** | | | | **Pool positivity** | | |
| --- | --- | --- | --- | --- | --- | --- | --- | --- |
| **Year** | **Month** | **Prev MLE** | **95% CI (MLE)** | **Prev Bayesian** | **95% CrI (Bayesian)** | **No. Pools** | **Pos. Pools** | **Pool +ve**  **(%)** |
| 2022 | Sep | 0.0053 | 0.0003 – 0.0231 | 0.0062 | 0.0005 – 0.0239 | 10 | 1 | 10.0 |
| 2022 | Oct | 0.0196 | 0.0078 – 0.0395 | 0.0197 | 0.0074 – 0.0404 | 18 | 6 | 33.3 |
| 2022 | Nov | 0.0219 | 0.0079 – 0.0469 | 0.0218 | 0.0074 – 0.0478 | 15 | 5 | 33.3 |
| 2022 | Dec | 0 | 0.0000 – 0.0232 | 0 | 0.0000 – 0.0235 | 6 | 0 | 0.0 |
| 2023 | Jan | 0 | 0.0000 – 0.0240 | 0 | 0.0000 – 0.0232 | 7 | 0 | 0.0 |
| 2023 | Feb | 0 | 0.0000 – 0.0663 | 0 | 0.0000 – 0.0644 | 3 | 0 | 0.0 |
| 2023 | Mar | 0.0041 | 0.0013 – 0.0094 | 0.0043 | 0.0014 – 0.0092 | 47 | 4 | 8.5 |
| 2023 | Apr | 0.0057 | 0.0031 – 0.0096 | 0.0057 | 0.0031 – 0.0097 | 97 | 12 | 12.4 |
| 2023 | May | 0.0091 | 0.0047 – 0.0155 | 0.0092 | 0.0049 – 0.0160 | 62 | 11 | 17.7 |
| 2023 | Jun | 0.0059 | 0.0018 – 0.0137 | 0.0062 | 0.0020 – 0.0145 | 36 | 4 | 11.1 |
| 2023 | Jul | 0.0025 | 0.0001 – 0.0111 | 0.003 | 0.0003 – 0.0110 | 26 | 1 | 3.8 |
| 2023 | Aug | 0.0177 | 0.0055 – 0.0407 | 0.0182 | 0.0065 – 0.0410 | 20 | 4 | 20.0 |
| 2023 | Sep | 0.0236 | 0.0059 – 0.0606 | 0.0245 | 0.0067 – 0.0607 | 10 | 3 | 30.0 |

**Monthly pool positivity and MLE infection prevalence estimate in *Cx. quinquefasciatus***

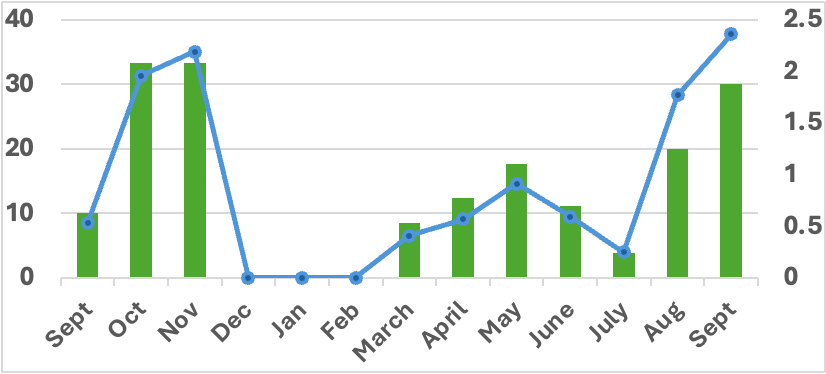

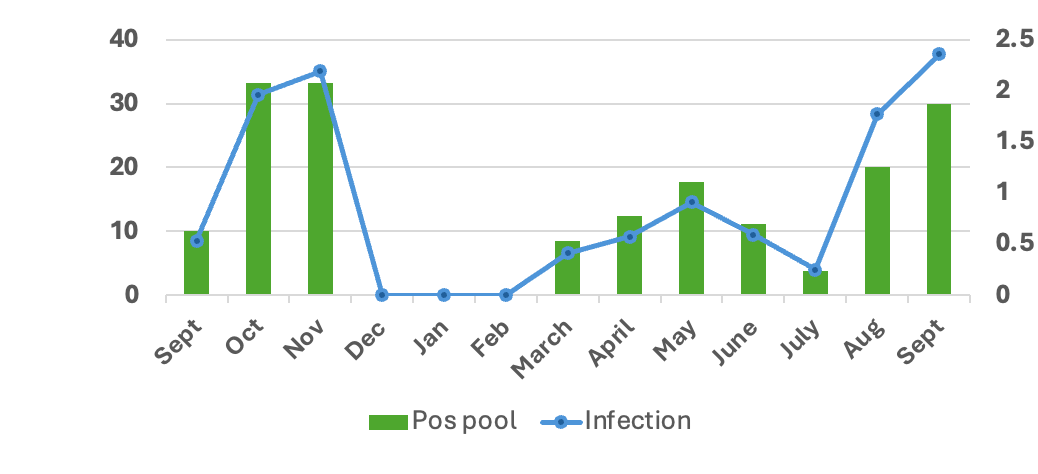

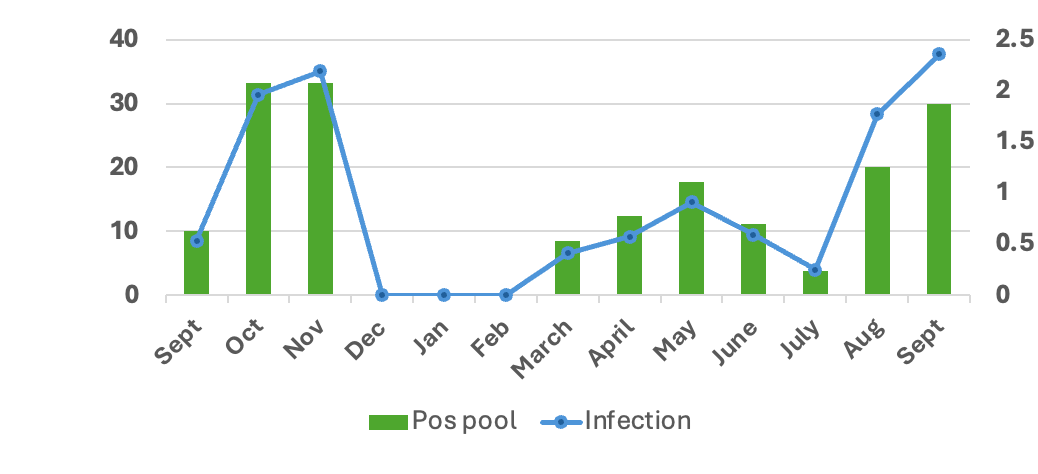

**Pool positivity**

**Infection prevalence (MLE)**

**S19. Table. Jhapa - Outputs from the PoolScreen analysis in R studio and pool positivity summaries**

|  |  | **Poolscreen analysis –infection prevalence** | | | | **Pool positivity** | | |
| --- | --- | --- | --- | --- | --- | --- | --- | --- |
| **Year** | **Month** | **Prev MLE** | **95% CI (MLE)** | **Prev Bayesian** | **95% CrI (Bayesian)** | **No. Pools** | **Pos. Pools** | **Pool +ve**  **(%)** |
| 2023 | Mar | 0.0043 | 0.0002 – 0.0186 | 0.0049 | 0.0005 – 0.0180 | 11 | 1 | 9.1 |
| 2023 | Apr | 0.0101 | 0.0040 – 0.0204 | 0.0103 | 0.0043 – 0.0209 | 33 | 6 | 18.2 |
| 2023 | May | 0.0146 | 0.0084 – 0.0232 | 0.0147 | 0.0085 – 0.0233 | 56 | 15 | 26.8 |
| 2023 | Jun | 0.0104 | 0.0045 – 0.0201 | 0.0108 | 0.0049 – 0.0203 | 37 | 7 | 18.9 |
| 2023 | Jul | 0.024 | 0.0145 – 0.0368 | 0.024 | 0.0146 – 0.0366 | 49 | 18 | 36.7 |
| 2023 | Aug | 0.0148 | 0.0088 – 0.0229 | 0.0156 | 0.0093 – 0.0236 | 63 | 17 | 27.0 |

**S20. Table. Kapilvastu - Outputs from the PoolScreen analysis in R studio and pool positivity summaries**

|  |  | **Poolscreen analysis –infection prevalence** | | | | **Pool positivity** | | |
| --- | --- | --- | --- | --- | --- | --- | --- | --- |
| **Year** | **Month** | **Prev MLE** | **95% CI (MLE)** | **Prev Bayesian** | **95% CrI (Bayesian)** | **No. Pools** | **Pos. Pools** | **Pool +ve**  **(%)** |
| 2023 | Mar | 0.019 | 0.0011 – 0.0828 | 0.0205 | 0.0018 – 0.0793 | 3 | 1 | 33.3 |
| 2023 | Apr | 0.0092 | 0.0033 – 0.0198 | 0.0092 | 0.0036 – 0.0194 | 27 | 5 | 18.5 |
| 2023 | May | 0.0052 | 0.0016 – 0.0121 | 0.0054 | 0.0018 – 0.0122 | 39 | 4 | 10.3 |
| 2023 | Jun | 0.0036 | 0.0006 – 0.0110 | 0.0038 | 0.0008 – 0.0107 | 27 | 2 | 7.4 |
| 2023 | Jul | 0.0051 | 0.0016 – 0.0117 | 0.0054 | 0.0016 – 0.0122 | 42 | 4 | 9.5 |
| 2023 | Aug | 0.0084 | 0.0033 – 0.0169 | 0.0084 | 0.0036 – 0.0173 | 40 | 6 | 15.0 |

**S21. Table. Epidemiological week–wise variation in LF pool positivity (%) and female *Culex quinquefasciatus* abundance in Dhading district (September 2022–September 2023)**

| **Epi week** | **Mosquitoes number** | **Number of pool** | **Mosquitoes number/Number of pool Ratio** | **Positive Pool number** | **Positive percentage** |
| --- | --- | --- | --- | --- | --- |
| 38-41 | 276 | 14 | 20 | 2 | 14.3 |
| 42-45 | 327 | 16 | 20 | 5 | 31.3 |
| 46-49 | 260 | 15 | 17 | 4 | 26.7 |
| 50-52 | 62 | 4 | 16 | 0 | 0.00 |
| 1-4 | 9 | 2 | 5 | 0 | 0.00 |
| 5-8 | 70 | 8 | 9 | 0 | 0.00 |
| 9-12 | 676 | 33 | 20 | 1 | 3.0 |
| 13-16 | 2093 | 91 | 23 | 11 | 12.1 |
| 17-20 | 1355 | 61 | 22 | 12 | 19.7 |
| 21-24 | 912 | 43 | 21 | 6 | 14.0 |
| 25-28 | 554 | 30 | 18 | 1 | 3.3 |
| 29-32 | 210 | 17 | 12 | 1 | 5.9 |
| 33-36 | 288 | 22 | 13 | 6 | 27.3 |
| 37-38 | 73 | 4 | 18 | 1 | 25.0 |

Epi-week wise distribution of LF Pool positive (Percentage) in Dhading District

Culex Quinquefasciatus female number

Pool positivity percentage

|  | Positive percentage |
| --- | --- |
| 38-41 | 14.2857 |
| 42-45 | 31.25 |
| 46-49 | 26.6667 |
| 50-52 | 0 |
| 1-4 | 0 |
| 5-8 | 0 |
| 9-12 | 3.0303 |
| 13-16 | 12.0879 |
| 17-20 | 19.6721 |
| 21-24 | 13.9535 |
| 25-28 | 3.33333 |
| 29-32 | 5.88235 |
| 33-36 | 27.2727 |
| 37-38 | 25 |

| Epi- week (SEP 2022 to Sep 2023) | Mosquitoes number |
| --- | --- |
| 38-41 | 276 |
| 42-45 | 327 |
| 46-49 | 260 |
| 50-52 | 62 |
| 1-4 | 9 |
| 5-8 | 70 |
| 9-12 | 676 |
| 13-16 | 2093 |
| 17-20 | 1355 |
| 21-24 | 912 |
| 25-28 | 554 |
| 29-32 | 210 |
| 33-36 | 288 |
| 37-38 | 73 |
